# A dollar-aware food-environment index and a 27-year trajectory typology: a measurement foundation for diet and childhood-obesity research in Mississippi, 1997–2024

**DOI:** 10.64898/2026.08.20.26360912

**Authors:** Sai Venkat Mandalapu, Sage Lefebvre, Erica D. Walker

## Abstract

**Background:** The retail food environment is a widely used exposure in behavioural-nutrition and obesity research. Grocery stores have declined across rural America while limited-assortment discount (“dollar”) stores have proliferated, yet standard food-environment indices typically exclude dollar stores. We developed a dollar-aware index, assessed how counting dollar stores changes the measured exposure, and derived a trajectory typology.

**Methods:** Using annual establishment data from Data Axle for all 878 Mississippi census tracts (1997–2024), we classified food retailers with code-based rules and computed the modified Retail Food Environment Index (mRFEI) in standard and dollar-aware forms. We fitted latent class tra-jectory models suited to the bounded index, tested their stability in seven sensitivity analyses, and related class membership to the Social Vulnerability Index (SVI) and urbanicity with multinomial regression.

**Results:** Grocery stores fell from 1,616 to 716 while dollar stores rose from 315 to 1,005, crossing in 2018. The indices ranked tracts almost identically (within-year Spearman ≥ 0.99), but where dollar stores and healthy outlets coexisted, counting dollar stores lowered the index by 4.5 points in 1997 and 9.1 in 2024, and tracts with only dollar-store retail, which the standard index cannot score, rose from 1 to 32. Six trajectory classes emerged: stable adequate (7.8% of tracts), steady decline (15.3%), early collapse (12.9%), late collapse (9.7%), persistently constrained (34.8%) and chronically low (19.6%); only 7.5% of children in classified tracts lived in stable-adequate tracts. Classes were stable across random starts and pandemic-year exclusion; collapse was robust, but its early–late timing split depended on tracts with few retailers. Chronically low and persistently-constrained tracts were predominantly urban (relative risk ratios 8.3 and 4.5), consistent with food swamps; persistently-constrained and steady-decline membership rose modestly with vulnerability (1.17 and 1.20 per 0.1 SVI), an association that was weaker with 2010 SVI; collapse classes had no cross-sectional social signature.

**Conclusions:** Counting dollar stores changes the level of a widely used food-environment index, and whether a growing set of tracts can be scored, rather than how tracts rank. A 27-year typology separates urban food swamps from tracts whose food environments collapsed and provides a time-varying exposure for diet and childhood-obesity research.

**Contributions to the literature:**

- Standard food-environment indices omit dollar stores; counting them barely changes how tracts rank, but it lowers index values where dollar stores and healthy outlets coexist and determines whether a growing set of tracts can be scored at all.
- We provide a 27-year, tract-level trajectory typology of a dollar-aware index, tested in seven sensitivity analyses, that distinguishes urban food swamps from communities whose food environments collapsed.
- Tracts whose food environments collapsed are indistinguishable from others on present-day social characteristics, illustrating what cross-sectional exposure measures miss.
- We document data issues that longitudinal food-environment studies using commercial data should check: vendor coverage expansions, store-size fields that shift with vendor coding, and code-based misclassification.

## 1. Background

The retail food environment, the mix of outlets from which a community obtains food, is one of the most widely used place-based exposures in behavioural-nutrition research [1,2]. Its appeal is mechanistic: the assortment, affordability and quality of nearby food are thought to shape diet quality, food security and, over time, weight and diet-related chronic disease [1,2]. Evidence linking local food environments to obesity is mixed and comes largely from cross-sectional studies of modest quality, although indices that summarise the overall mix of outlets have been associated with obesity more consistently than counts of single outlet types [3]. Food deserts, characterised by limited access to healthy foods [4], and food swamps, where energy-dense, nutrient-poor outlets saturate an area, are two widely recognised food environments, with food-swamp conditions predicting obesity prevalence at least as strongly as food deserts [5]. Characterising either exposure therefore requires attention to both the availability of food outlets (how many there are and how near) and the composition of local food retail (the mix of healthy and less-healthy outlet types).

The food retail landscape in the United States has shifted substantially, particularly in rural and small-town communities [6,7]. Over the past two decades, independent grocery stores have closed in large numbers [8,9], while limited-assortment discount general-merchandise retailers, colloquially “dollar stores”, have expanded rapidly [10,6]. Dollar stores are small-format stores, dominated by a few national chains (Dollar General, Dollar Tree and Family Dollar), that sell low-priced, mostly shelf-stable foods alongside household goods [9]; they compete with other formats in ways that have been compared with the hard discounters found in other countries [11], but they carry a narrower food range with little fresh food [12,13]. Their growth has been concentrated in low-income and rural communities and in communities of color [10,6,7], and dollar-store entry has been followed by the contraction or exit of independent grocers, particularly in rural areas [8,9], although one dynamic model of entry found it to be followed by the exit of other small-format stores rather than supermarkets [11], and losses of food access after entry appear concentrated in urban neighbourhoods with a single grocery store [14]. Store audits describe shelves dominated by energy-dense, highly processed items with little or no fresh produce [12,13,15], even though many shoppers buy food there regularly [16,6]. Evidence on diet is more mixed: foods bought at dollar stores are generally less healthy than those bought elsewhere, but the overall healthfulness of household food purchases was similar between households that did and did not shop there, consistent with compensating purchases at other outlets [17], and some dollar stores offer fresh produce of comparable quality at lower prices [18]. Dollar stores are also widely authorised to accept Supplemental Nutrition Assistance Program (SNAP) benefits [13], whereas authorisation for the Special Supplemental Nutrition Program for Women, Infants, and Children (WIC) requires vendors to stock minimum varieties and quantities of specified foods [19]; where dollar stores replace other retailers, WIC families may therefore lose places to redeem benefits faster than SNAP families.

How the retail food environment is measured determines the exposures used in diet and obesity research, yet the decisions about which outlets count as healthy are rarely examined, even though the classification of outlets in commercial databases is known to be imperfect [20,21,22]. The modified Retail Food Environment Index (mRFEI), one of the most widely used measures and an adaptation of an earlier ratio-based index [23], expresses the proportion of food retailers classified as “healthy” (supermarkets, larger grocery stores and supercentres) among all healthy and “less-healthy” outlets (convenience stores, fast-food restaurants and small grocery stores) [24]. Dollar stores are not among the outlet types the index counts [24], because they are neither full-service grocers nor conventional convenience stores or restaurants. They are also inconsistently represented in the commercial and administrative business databases used to construct food-environment exposures, where classification practices and industry codes have changed repeatedly over time [20,21]. Whether a proliferating limited-assortment format is counted among less-healthy outlets is not a property of the data but a decision by the analyst, and as the format expands the decision may matter more. This raises a measurement question: does counting dollar stores change how tracts rank, their index values, or whether they can be measured at all, and how does its effect evolve as dollar stores proliferate? Neither version should be regarded as “ground truth”. Classifying dollar stores as less healthy is a defensible simplification that reflects their typical inventory, but they are not equivalent to fast-food outlets, and some now stock fresh produce [18].

A second limitation of food-environment exposures is that they are typically measured cross-sectionally [3,2]: in a review of 432 food-environment studies, only 13.9% used a longitudinal design [25]. A single snapshot misses how food retail, diet and health evolve over years, and studies linking time-varying food-retail measures to diet have produced more mixed associations than the cross-sectional literature that shaped food-desert policy [26]. Communities also do not change uniformly: some lose food retail abruptly, others gradually, and others remain persistently underserved. Time-varying summaries of how small areas diverge over decades, suitable for life-course exposure modelling, remain uncommon, although latent class trajectory models offer one established way to construct them [27,28]. Deriving such a summary is a further goal.

We focus on Mississippi because it is a priority setting for diet-related disease: more than 40% of adults had obesity in 2024, one of only two states at that level [29]; 25.0% of youth aged 6 to 17 years had obesity in 2022–2023, the highest of any state [30]; and nearly one in five residents lives below the poverty line [31]. In this setting we pursue four aims: (1) construct a longitudinal, dollar-aware food-environment exposure and quantify how the measured environment changes when dollar stores are counted; (2) characterise the 27-year trajectory of food-retail composition; (3) derive latent trajectories of tract-level food-retail adequacy as a time-varying exposure typology and test their robustness; and (4) examine how those trajectories relate to social vulnerability, urbanicity and spatial structure. Because food-environment measures are routinely used as exposures in studies of diet, food security and diet-related disease, including childhood obesity, understanding how a changing retail landscape alters those measures is a prerequisite for interpreting the studies that rely on them. Evidence linking dollar stores specifically to children’s weight is limited: in neighbouring Arkansas, the presence of dollar stores near children’s homes was not associated with higher body mass index [32]. Accurate, time-varying exposure measures are a prerequisite for testing such associations, and the measures developed here will serve as exposures in a planned longitudinal study of childhood obesity in Mississippi.

## 2. Methods

### 2.1 Data source and study area

Establishment-level records were obtained from Data Axle (formerly Infogroup, whose database was previously known as InfoUSA), a commercial business database compiled from public records, directories and direct verification. Commercial establishment lists are widely used to locate and classify food outlets; against field observation they show moderate to high agreement, with InfoUSA among the more accurate sources, although small and independent outlets are more often missed [20,21,33,22]. We obtained the complete annual Mississippi files for 1997–2024 (28 files). Each record includes the establishment name, address, coordinates, primary and secondary Standard Industrial Classification (SIC) and North American Industry Classification System (NAICS) codes, location employee size, a unique establishment identifier and, where applicable, a corporate-parent identifier. An establishment was counted as operating in a year if it appeared in that year’s file. Coordinates were missing for a negligible number of records, which could not be assigned to a tract (for example, one grocery store in 2003). The study area comprised all 878 census tracts in Mississippi; census tracts are small statistical areas defined by the US Census Bureau, generally home to 1,200–8,000 people [34].

### 2.2 Retail classification and validation

Food retailers were assigned to five categories: grocery stores, supercentres, convenience stores (including gas stations with a store), dollar stores and fast-food outlets. Because industry codes classify dollar stores inconsistently across SIC and NAICS vintages (Additional file 1: Table S1), we identified dollar stores from three signals in each record: establishment name, corporate-parent identifier and NAICS code. Names were matched against the national and regional chains operating in Mississippi (Dollar General, including DG Market and pOpshelf; Family Dollar; Dollar Tree; Fred’s discount stores, excluding Fred’s pharmacies; Big Lots; Five Below; Ollie’s Bargain Outlet; and Super 10) and a screened list of local dollar-store chains with more than two Mississippi locations. Big Lots and Ollie’s Bargain Outlet are larger closeout formats but were included because both sell packaged foods: food and beverages accounted for 12–13% of Big Lots’ net sales in fiscal 2020–2021 [35], and Ollie’s stocks packaged foods including canned goods, cereal and snacks [36]; excluding both reduced dollar-store establishment-years by 2.3%; Dirt Cheap, a regional closeout chain, was excluded; corporate-parent identifiers, which are numeric and therefore unaffected by spelling variation, recovered chain stores whose names were recorded inconsistently (for example, Fred’s stores in 2004–2012). Of the 18,582 dollar-store establishment-years, 76.3% matched on both name and parent identifier, 18.5% on name alone and 5.2% on parent identifier alone; 113 records (0.6%) matched more than one chain and were assigned to a single chain by a fixed labelling rule, which does not affect dollar-store counts. Name and parent identifier established chain membership, and the NAICS code served as a confirming signal: 98.2% of identified dollar stores carried a variety- or general-merchandise NAICS code. The NAICS signal was not used on its own, because the same codes also cover supercentres and department and apparel stores (14,284 further records), and because the contemporary dollar-store codes alone would have missed 77–98% of dollar stores before 2006 and about 3–6% by the early 2020s (Additional file 1: Table S1). Annual counts for Dollar General, the largest chain, were within 0.8–7.6% of the Mississippi store totals the company reported in its Form 10-K filings for 2017–2024 (mean absolute difference 4.6%), a slight, consistent undercount with no evidence of overcounting [37] (Additional file 1: Table S2).

Grocery stores, supercentres and convenience stores were classified by name and industry code, with explicit exclusion of non-food establishments carrying food-retail codes (florists, pharmacies, restaurants, used-car dealers, tobacco retailers, frozen-food wholesalers and hardware stores). Supercentres generated multiple records at a single location in later years; only the principal record per location was retained. The SIC-to-NAICS transition around 2008 introduced discontinuities in grocery and convenience counts that reflected re-coding rather than retail change; these were resolved using exact NAICS sub-codes. Fast-food outlets were limited-service restaurants identified by name and industry code, excluding outlets located inside other stores. The categories were mutually exclusive except for one establishment-year counted as both a grocery and a convenience store. The standard mRFEI counts grocery stores with three or fewer employees as less healthy [24]. We counted all grocery stores as healthy because the employee-size field shifted with vendor coding during the study period (the share of grocery records with three or fewer employees rose from 41% in 2012 to 54% in 2013–2015 and fell to 46% in 2016), which would introduce artificial change into a longitudinal measure; the size-based definition is reported as a sensitivity analysis.

Classification was rule-based and implemented in code rather than assigned by hand, so it is fully reproducible. Rather than inter-coder reliability, we therefore assessed the output for systematic misclassification. We identified four sources: electric-vehicle charging stations carrying gas-station codes from 2013 onward (652 records; 1.0% of convenience records), convenience-format stores carrying vendor grocery codes (1,024 records; 3.3% of grocery records), café listings within warehouse clubs classified as supercentres (14 records) and grocery-format Walmart Neighborhood Markets classified as supercentres (103 records; because both categories count as healthy, the index is unaffected) (Additional file 1: Table S3). Their effect on the index and on the trajectory classes was assessed in a sensitivity analysis.

### 2.3 The dollar-aware food-environment index

We computed the mRFEI for each census tract and year as the percentage of healthy retailers (grocery stores and supercentres) among all healthy and less-healthy retailers, where less-healthy retailers comprise convenience stores and fast-food outlets [24]. The dollar-aware mRFEI additionally counts dollar stores as less-healthy retailers, adding them to the denominator and leaving the numerator unchanged. Both indices range from 0 to 100, with higher values indicating a healthier outlet mix, and are undefined in tract-years with no retailer in their denominator. Counting dollar stores as less healthy is a simplification: assortments vary across stores and over time, and some dollar stores offer fresh produce [18]; we return to this assumption in the Discussion.

Because the two indices differ only in how dollar stores are classified, their difference isolates that single decision. A difference can arise only in tract-years containing both a dollar store and a healthy outlet: it is zero where a tract has no dollar store, and also where it has no healthy outlet, because both indices are then 0. We therefore report (i) the mean difference among all tract-years both indices can score, (ii) the mean difference among tract-years in which a difference is possible, and (iii) the number of tract-years whose only food retailers were dollar stores, which the standard index cannot score and the dollar-aware index scores as 0. We also report the within-year rank correlation (Spearman) between the two indices. Neither index is treated as ground truth (see Background).

### 2.4 Census tract harmonisation, social and demographic data

Census-tract boundaries changed after the 2000, 2010 and 2020 censuses, so we assigned every establishment in every year to 2020 census tracts (TIGER/Line 2022) [38] by point-in-polygon matching of its coordinates. Tract characteristics were taken from the CDC/ATSDR Social Vulnerability Index (SVI) 2022 (Mississippi, state-ranked), a composite percentile ranking on 16 social factors [39,40], and from American Community Survey (ACS) 2018–2022 five-year estimates [31] of population, percentage Black, percentage in poverty, median household income and percentage younger than 18 years; the number of children in each tract was the population multiplied by the percentage younger than 18 years. As an earlier-period measure, we mapped SVI 2010 (Mississippi, state-ranked, 2010 tracts) [41] to 2020 tracts using the Census Bureau’s 2020–2010 tract relationship file [42], weighting by shared land area (872 of 878 tracts matched). A tract was classified as urban if its 2020 population-weighted centroid [43] fell within a 2020 Census urban area [38].

### 2.5 Statistical analysis

We described the statewide trajectory of each retail category and estimated compound annual growth rates by regressing the natural logarithm of statewide counts on year. Because the coverage and categorisation of commercial business data can change over time [44], we examined year-on-year change in the total number of Mississippi records in each annual file and in each retail category to distinguish retail change from changes in vendor coverage.

To derive a time-varying exposure typology, we fitted group-based trajectory models [45] to the tract-level dollar-aware index for 1997–2024 as latent class mixed models with cubic class-specific trajectories and no within-class random effects (lcmm package [46]); without random effects, classes describe shared mean shapes. Because the index is bounded at 0 and 100 and 30.4% of modelled tract-years were exactly 0, the primary model linked the index to the latent trajectory through a monotone spline transformation with five equidistant knots, which accommodates floor and ceiling effects [46]; a beta-link alternative did not converge. A linear (Gaussian) specification is reported as a sensitivity analysis (Additional file 1). Tract-years with no food retailer, for which the index is undefined, were treated as missing. Tracts with at least one food retailer in at least 15 of the 28 years were modelled (n = 817); the remaining 61 tracts, 17 of which never had a food retailer, could not support a trajectory and are described separately.

Models with one to eight classes were fitted from 24 random starts each and the best start was iterated to convergence. The number of classes was chosen by considering the Bayesian Information Criterion (BIC), the sample-size-adjusted BIC, relative entropy, the minimum average posterior probability of assignment (APPA; adequate *≥*0.7) and occupancy odds ratio (OCC; adequate *≥*5) [45], the size of the smallest class, the stability of classes across adjacent solutions and their sub-stantive distinctness [27,28]. The classes were named by the authors to describe the shape of their trajectories; the names are descriptive and do not by themselves imply a food desert or a food swamp, which we consider in the Discussion. Model fit was assessed by comparing model-predicted class trajectories with observed class means.

We assessed the stability of the selected solution by refitting it (i) from independent random starts; (ii) excluding tracts averaging fewer than three food retailers per year, the level below which a single opening or closure moves the index substantially (year-on-year changes of 50 points or more occurred in 11–12% of transitions from tract-years with one or two retailers but in under 2% from tract-years with three or more); (iii) treating tract-years with fewer than three retailers as missing and retaining tracts with at least 15 such years; (iv) excluding 2020 and 2021; (v) with the size-based small-grocery definition; (vi) after correcting the misclassifications described above; and (vii) with the linear specification. Agreement with the main solution was summarised by the adjusted Rand index (ARI) [47] and by the percentage of tracts retaining their class after matching classes by majority overlap.

We related class membership to tract characteristics with multinomial logistic regression (reference class: stable adequate), fitting one model with the overall SVI and urbanicity and a second with the SVI components (percentage Black, percentage in poverty and median household income) and urbanicity, because the overall index is constructed partly from these components. Relative risk ratios (RRR) are expressed per 0.1 of SVI (10 percentile points), per 10 percentage points and per US$10,000. Because class membership is spatially clustered, we report 95% confidence intervals from a county cluster bootstrap [48] (1,000 resamples of Mississippi’s 82 counties; percentile intervals) as the primary inference, alongside model-based intervals. The SVI and ACS measures describe 2018–2022, the end of the study period, and could partly reflect the food-environment change under study, so these associations are cross-sectional; we repeated the SVI model with SVI 2010 as a sensitivity analysis.

To characterise access, we computed the distance from each tract’s 2020 population-weighted centroid to the nearest operating grocery store, and to the nearest grocery store or supercentre, in 2024. We assessed spatial clustering of class membership with join-count statistics [49] under first-order queen contiguity and of index values with the Getis-Ord Gi* statistic [50] (Additional file 1: Supplementary Methods S5).

Analyses were conducted in ArcGIS Pro and R (lcmm, sf, spdep, nnet and ggplot2 packages). This study is reported in accordance with the STROBE guidelines (Additional file 2) and the GRoLTS checklist for latent trajectory studies [51] (Additional file 3).

## 3. Results

### 3.1 Food-retail composition and the grocery–dollar crossover

Statewide, the number of operating grocery stores fell from 1,616 in 1997 to 716 in 2024 (*−*55.7%; *−*3.0% per year), while dollar stores rose from 315 to 1,005 (+219%; +4.1% per year); the two crossed in 2018, and the dollar-to-grocery ratio rose from 0.19 to 1.40 (Fig. 1a; Additional file 1: Table S4). Convenience stores, the most numerous category, changed little (*−*5.8%), fast-food outlets increased by 47.7%, and supercentres grew from 12 to 91. The total number of records in the annual files rose abruptly in 2001 (+17.3%) and in 2012–2013 (+9.0% and +8.2%) (Additional file 1: Table S5; Fig. S1a). In 2012 every retail category rose, including grocery stores (+13.4%), which otherwise rose in only three years and by at most 3.2%; 18.4% of that year’s grocery records were new to the database, against a median of 8.0% (range 1.3–14.4%) in other years, and fewer than half of these (41.4%) were still listed as grocery stores in 2016. We therefore interpret the 2012 rise as an expansion of vendor coverage rather than retail change; because the mRFEI is a ratio, coverage changes that affect all categories proportionally largely cancel. The database also contracted between 2015 and 2020, but grocery stores continued to fall as a share of all records, so the grocery decline is not an artefact of coverage. In 2020, convenience stores (*−*6.8%), grocery stores (*−*6.6%) and dollar stores (*−*6.1%) all declined, which may reflect pandemic-related closures, disrupted vendor verification, or both; the trajectory classes were largely unchanged when 2020 and 2021 were excluded (Section 3.4).

**Figure 1.**
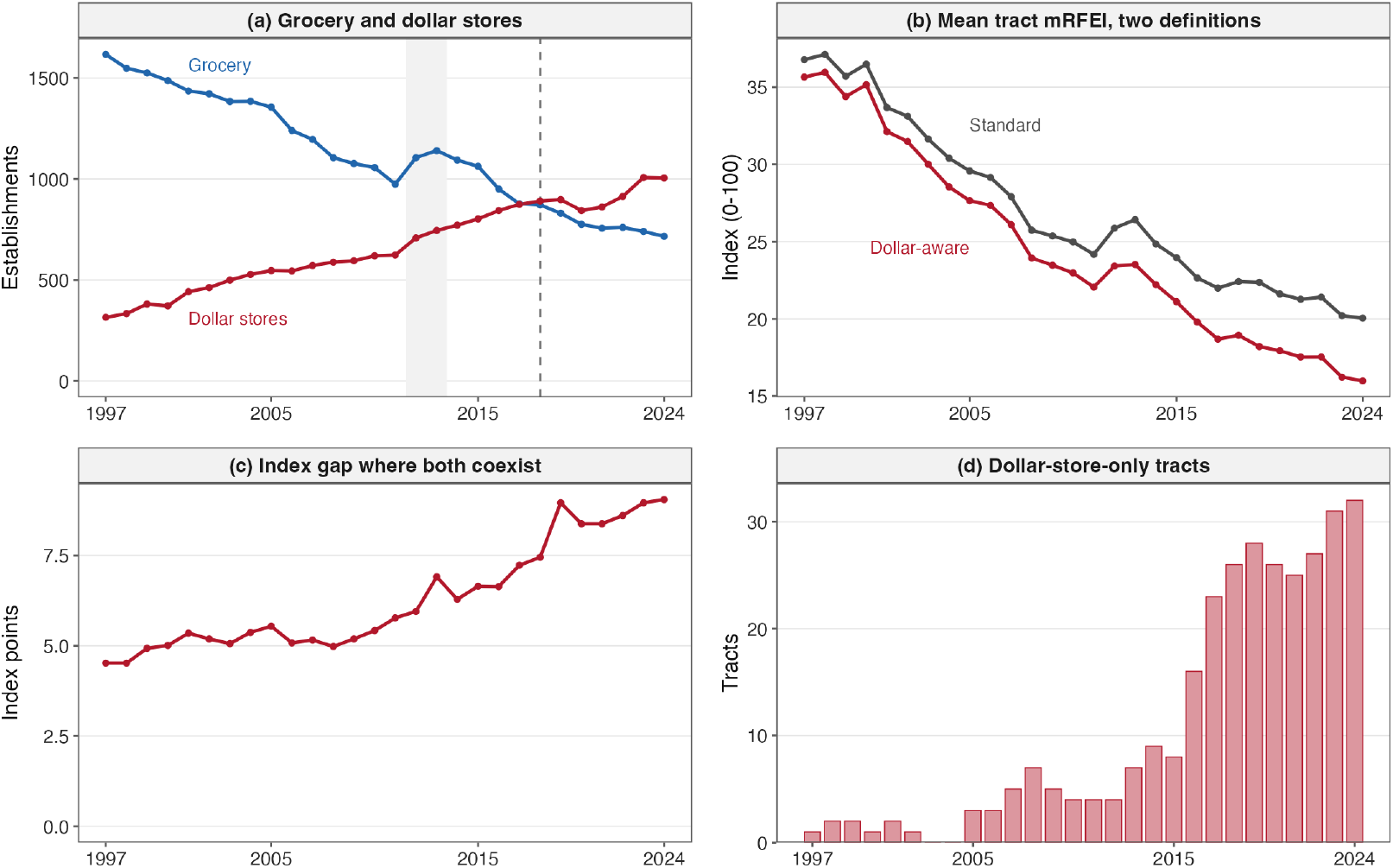
Food retail and the retail food environment index in Mississippi, 1997–2024. (a) Statewide grocery and dollar stores; dashed line, 2018, the first year dollar stores outnumbered grocery stores; shaded, 2012–2013, a database-wide expansion of vendor coverage (all outlet types in Additional file 1: Table S4). (b) Mean tract modified Retail Food Environment Index (mRFEI) under the standard and dollar-aware definitions, among tracts both can score. (c) Mean difference between the two in tract-years where dollar stores and healthy outlets coexist. (d) Tracts whose only food retailers were dollar stores, which the standard index cannot score. Source: Data Axle.

### 3.2 How counting dollar stores changes the measured exposure

The standard and dollar-aware indices share a numerator and differ only in whether dollar stores enter the denominator. They ranked tracts almost identically within each year (Spearman correlation 0.991–0.996; fewer than 3.5% of tracts moved by more than 10 percentile points; Additional file 1: Table S6). The indices differed in level. Across all tract-years both could score, the mean difference rose from 1.1 points in 1997 to 4.1 points in 2024 (Fig. 1b), but this average includes the many tract-years in which a difference is impossible. Where dollar stores and healthy outlets coexisted, counting dollar stores lowered the index by 4.5 points in 1997 and 9.1 points in 2024 (medians 3.1 and 4.3; Fig. 1c). The share of scored tracts containing a dollar store rose from 27.4% to 69.9%, and among these the share with no healthy outlet, in which both indices are 0, rose from 9.2% to 35.7%. Where a difference was possible, it was largest in the least socially vulnerable quartile (16.4 points in 2024) and smallest in the most vulnerable quartile (5.5 points; Additional file 1: Fig. S1b,c). The indices also differed in which tracts could be measured: tracts whose only food retailers were dollar stores, scored 0 by the dollar-aware index but undefined by the standard index, rose from 1 in 1997 to 32 in 2024 (302 tract-years in 72 tracts; Fig. 1d). Of the 72 tracts whose only food retailers were dollar stores in at least one year, 32 were in the least vulnerable SVI quartile and 8 in the most.

### 3.3 A six-class trajectory typology

Of the 878 tracts, 817 had at least one food retailer in at least 15 of the 28 years and were modelled. In these tracts the median number of food retailers per tract-year was 5 (interquartile range 3–10), 23.0% of tract-years with any retailer had one or two, and 3.7% of tract-years had none and were treated as missing; 636 tracts had a food retailer in all 28 years. The 61 tracts not modelled had a median of 5 years with any food retailer and at most 7 retailers in any year; 17 never had one.

BIC declined with every additional class up to eight in both the spline-link and linear models, with diminishing improvements (0.62%, 0.42%, 0.38% and 0.31% for five to eight classes in the spline-link model; Additional file 1: Table S7; Fig. S2a). We selected six classes because six was the smallest solution that separated early from late collapse (the five-class solution merged them).

The seven-class solution split a small class from the persistently-constrained class (44 tracts, with 11 from the chronically low class) while retaining 93–98% of each other class, and the eight-class solution added a further mixed class of low-scoring tracts (Additional file 1: Table S7). The six-class solution had relative entropy 0.964, class-specific APPA of 0.953–0.983 and OCC of at least 80.7, and its smallest class held 7.8% of tracts (Additional file 1: Table S8). The spline-link model fitted far better than the linear specification (BIC 175,077 versus 189,145), and its predicted trajectories tracked observed class means closely (mean absolute difference 0.9–3.4 index points; Fig. 2); the largest single-year deviation was for the early-collapse class in 2002 (observed 44.3, predicted 29.3). The classes describe distinct exposure histories (Fig. 2; Table 1). A stable adequate class (64 tracts, 7.8%) held a high index throughout (observed class mean 67.6 in 1997 and 56.8 in 2024). A steady decline class (125 tracts, 15.3%) eroded gradually (47.9 to 29.1). Two classes shared a high-to-floor shape but differed in timing: the model-predicted trajectory of an early collapse class (105 tracts, 12.9%) fell below 20 by 2005 (47.4 to 4.3), whereas that of a late collapse class (79 tracts, 9.7%) declined through the 2000s and fell below 20 by 2013 (64.7 to 2.1). A persistently constrained class (284 tracts, 34.8%), the largest, remained low with slow erosion (23.7 to 14.8), and a chronically low class (160 tracts, 19.6%) stayed near the floor throughout (9.4 to 3.3).

**Table 1.** Characteristics of the six food-environment trajectory classes, Mississippi census tracts.

| Class | Tracts, n (%) | Children, n (%) | SVI | Black, % | Poverty, % | Income, \$ | Urban, % | Grocery, km | mRFEI 1997 | mRFEI 2024 |
|---|---|---|---|---|---|---|---|---|---|---|
| Stable adequate | 64 (7.8) | 49,109 (7.5) | 0.42 | 34.6 | 18.0 | 55,125 | 18.8 | 5.0 | 67.6 | 56.8 |
| Steady decline | 125 (15.3) | 102,793 (15.8) | 0.57 | 44.0 | 22.5 | 46,271 | 28.8 | 4.4 | 47.9 | 29.1 |
| Early collapse | 105 (12.9) | 79,592 (12.2) | 0.49 | 38.2 | 18.7 | 53,073 | 32.4 | 6.2 | 47.4 | 4.3 |
| Late collapse | 79 (9.7) | 60,143 (9.2) | 0.46 | 36.7 | 19.7 | 53,987 | 21.5 | 7.9 | 64.7 | 2.1 |
| Persistently constrained | 284 (34.8) | 238,956 (36.7) | 0.58 | 42.3 | 21.0 | 51,023 | 55.3 | 2.6 | 23.7 | 14.8 |
| Chronically low | 160 (19.6) | 121,271 (18.6) | 0.42 | 35.6 | 17.5 | 60,922 | 65.0 | 3.8 | 9.4 | 3.3 |

*Note:* Classes from a six-class latent class trajectory model of the dollar-aware mRFEI, 1997–2024, with a spline link (n = 817 tracts). % of tracts is the share of the 817 classified tracts; children are tract population younger than 18 years (American Community Survey 2018–2022), as a share of the 651,864 children in classified tracts. SVI is the class mean of the CDC/ATSDR Social Vulnerability Index 2022 overall percentile (0–1; higher = more vulnerable). Black, poverty and income are class means of tract percentage Black, percentage in poverty and median household income; urban is the share of tracts whose population-weighted centroid lies in a 2020 Census urban area. Grocery is the mean straight-line distance from the tract population-weighted centroid to the nearest grocery store in 2024. mRFEI 1997 and 2024 are observed class means of the dollar-aware index (0–100). The corresponding table for the linear specification is in Additional file 1.

**Figure 2.**
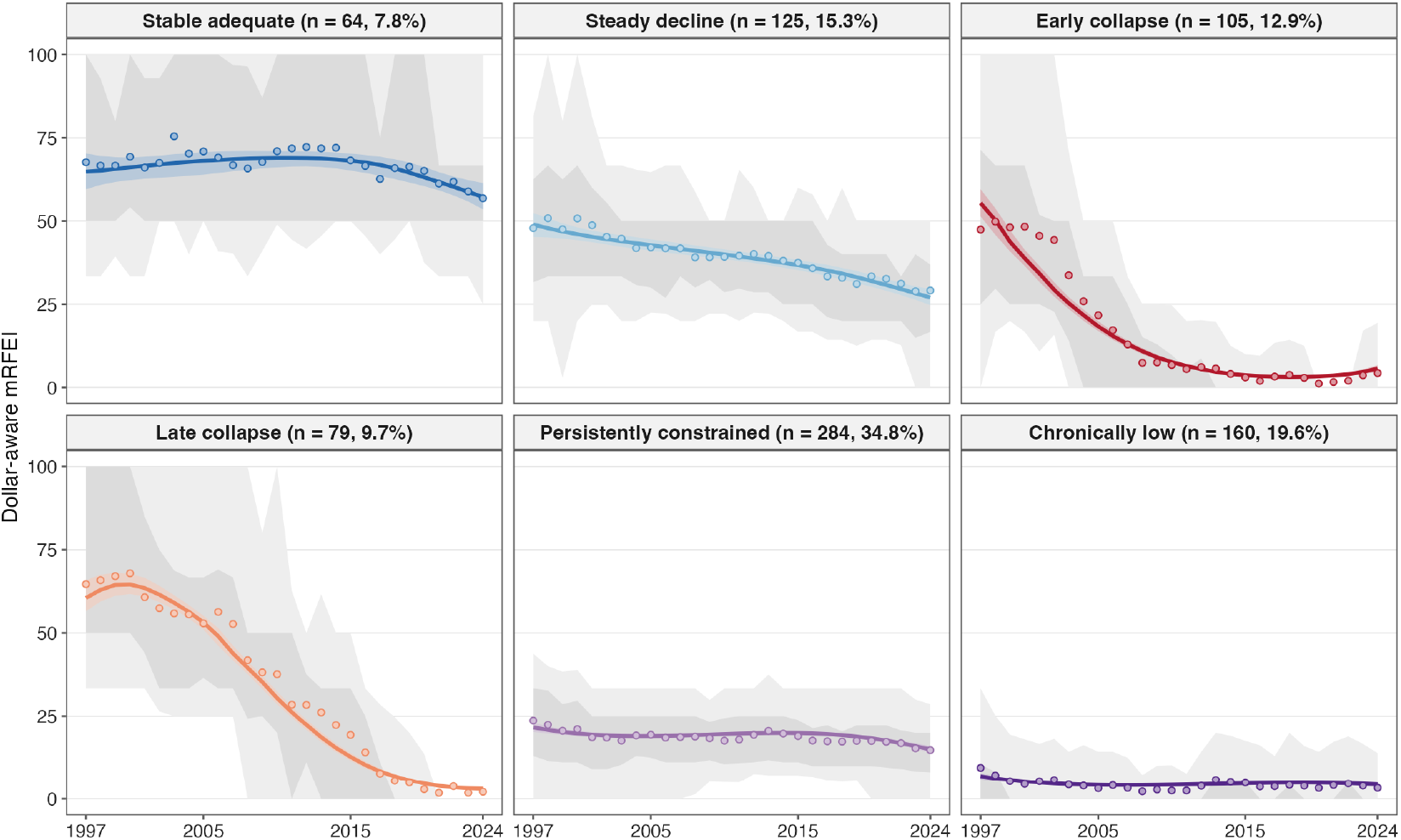
Trajectories of the dollar-aware mRFEI, 1997–2024, for six classes of Mississippi census tracts (latent class model with a spline link; 817 tracts). Grey bands, 10th–90th (light) and 25th–75th (dark) percentiles of tract values in each year; points, observed class mean; coloured line and band, model-predicted class trajectory with 95% interval. Years with no food retailer are treated as missing. Source: Data Axle.

### 3.4 Robustness of the typology

An independent refit from new random starts reproduced the six classes exactly (ARI 1.00), and the classification changed little when 2020 and 2021 were excluded (ARI 0.84; 91.8% of tracts retained their class) or when the misclassifications identified in the classification review were corrected (ARI 0.88; 94.5%) (Table 2). Correcting those misclassifications changed 5.7% of tract-year index values, lowering them by 0.75 points on average, consistent with the uncorrected index slightly overstating healthy access. Agreement with the linear specification was moderate (ARI 0.50; 74.4%; Additional file 1: Figs. S3–S4): the six trajectory shapes matched one-to-one, and most disagreements involved tracts that the linear model placed in the chronically low or persistently-constrained classes, or the timing of collapse. Excluding tracts with few retailers reduced agreement (ARI 0.53 when tracts averaging fewer than three retailers were excluded; 0.43 when tract-years with fewer than three retailers were set to missing). In both analyses the stable-adequate, steady-decline, persistently-constrained and chronically low classes largely persisted (71–100% of tracts retained) and a collapse class remained, but early and late collapse were no longer separated: one merged into the other (Additional file 1: Table S9). The existence of collapse is therefore robust, whereas its division by timing depends on tracts with few retailers, in which a single closure moves the index substantially. Counting grocery stores with three or fewer employees as less healthy lowered the index by 12.8 points on average and changed the classification substantially (ARI 0.20; 53.5%).

**Table 2.** Stability of the six-class solution across sensitivity analyses.

| Analysis | Spline link (primary): ARI | Spline link: % retaining class | Linear: ARI | Linear: % retaining class |
|---|---|---|---|---|
| Independent refit from new random starts | 1.00 | 100.0 | 1.00 | 100.0 |
| Excluding 2020 and 2021 | 0.84 | 91.8 | 0.90 | 93.6 |
| Correcting identified misclassifications | 0.88 | 94.5 | 0.84 | 93.5 |
| Excluding tracts averaging fewer than three food retailers per year | 0.53 | 82.0 | 0.81 | 89.7 |
| Tract-years with fewer than three food retailers set to missing | 0.43 | 76.1 | 0.58 | 79.6 |
| Grocery stores with three or fewer employees counted as less healthy | 0.20 | 53.5 | 0.20 | 55.3 |
| Agreement between the spline-link and linear specifications | 0.50 | 74.4 | 0.50 | 74.4 |

*Note:* ARI, adjusted Rand index (1 = identical classification). Each analysis was compared with the six-class solution of the same specification; % of tracts retaining class is computed after matching classes by majority overlap. Excluding tracts or tract-years reduces the sample (595 and 627 tracts). Retention by class is in Additional file 1: Table S9.

Of the 651,864 children living in the 817 classified tracts, 49,109 (7.5%) lived in stable-adequate tracts; the remainder lived in tracts whose index declined, collapsed or remained low throughout (Table 1). Including the 39,212 children in the 61 unclassified tracts lowers the stable-adequate share to 7.1%. We report these figures as a description of the exposure distribution, not a risk estimate.

### 3.5 Social patterning of the typology

Class membership was patterned more consistently by urbanicity than by social vulnerability (Fig. 3b,c; Table 3). Relative to the stable-adequate class, chronically low and persistently-constrained membership was strongly associated with urban location (RRR 8.33, 95% CI 4.34–26.17; and 4.49, 2.18–14.79). Persistently-constrained and steady-decline membership rose modestly with vulner-ability (RRR per 0.1 SVI 1.17, 1.06–1.31; and 1.20, 1.08–1.33). The collapse classes showed no significant association with either SVI or urbanicity. In the components model, percentage Black, percentage in poverty and median household income were not independently associated with class membership, whereas the urban associations persisted (Additional file 1: Table S10). With 2010 SVI, which correlated 0.71 with 2022 SVI, the vulnerability associations for the persistently-constrained and steady-decline classes weakened (RRR 1.05, 0.94–1.16; and 1.10, 0.97–1.24) and chronically low membership was associated with lower vulnerability (RRR 0.87, 0.76–0.98), whereas the urban associations were unchanged (RRR 8.54 and 5.20). In the linear specification, the vulnerability associations were stronger and persisted with 2010 SVI (Additional file 1: Table S10).

**Table 3.** Multinomial logistic regression of trajectory-class membership on social vulnerability and urbanicity.

| Trajectory class | SVI, per 0.1:<br>RRR<br>(cluster-bootstrap 95% CI) | Model-based<br>95% CI | Urban vs rural:<br>RRR<br>(cluster-bootstrap 95% CI) | Model-based<br>95% CI |
|---|---|---|---|---|
| Steady decline | 1.20 (1.08–1.33)* | 1.07–1.35 | 1.42 (0.72–4.65) | 0.67–3.01 |
| Early collapse | 1.08 (0.96–1.21) | 0.96–1.21 | 1.92 (0.92–5.28) | 0.90–4.09 |
| Late collapse | 1.06 (0.93–1.19) | 0.93–1.19 | 1.12 (0.38–4.59) | 0.49–2.59 |
| Persistently constrained | 1.17 (1.06–1.31)* | 1.06–1.30 | 4.49 (2.18–14.79)* | 2.27–8.85 |
| Chronically low | 0.95 (0.85–1.07) | 0.85–1.06 | 8.33 (4.34–26.17)* | 4.07–17.03 |

*Note:* Relative risk ratios (RRR) from a single multinomial logistic model of class membership on the CDC/ATSDR Social Vulnerability Index 2022 overall percentile (per 0.1, i.e. 10 percentile points) and urban location (tract population-weighted centroid in a 2020 Census urban area); reference class stable adequate; n = 816 tracts with a valid SVI. Primary 95% confidence intervals are percentile intervals from a county cluster bootstrap (1,000 resamples of 82 counties); model-based intervals are shown for comparison. \* cluster-bootstrap interval excludes 1. Associations are cross-sectional. Components models, models using 2010 SVI and the linear specification are in Additional file 1: Table S10.

**Figure 3.**
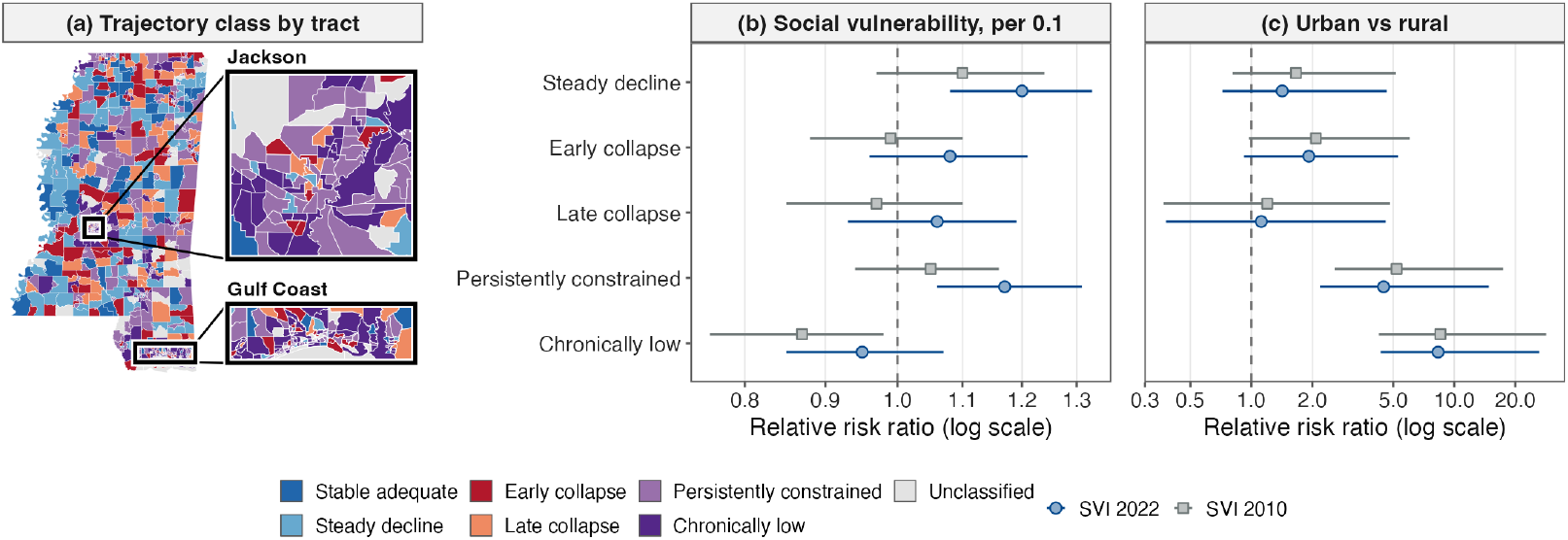
Geography and tract correlates of the trajectory classes. (a) Class of each Mississippi census tract, 1997–2024 (2020 boundaries), with the Jackson metropolitan area and the Gulf Coast enlarged at right (boxes mark their locations); unclassified, 61 tracts with a food retailer in fewer than 15 of 28 years. (b, c) Relative risk ratios from multinomial models with stable adequate as reference, for (b) social vulnerability (per 0.1, i.e. 10 percentile points) and (c) urban location (2020 urban area, population-weighted tract centroid), using 2022 SVI or, as a sensitivity analysis, 2010 SVI mapped to 2020 tracts; bars, 95% intervals from a county cluster bootstrap (1,000 replicates). Associations are cross-sectional. Source: Data Axle; CDC/ATSDR SVI 2010 and 2022; US Census Bureau TIGER/Line 2022.

### 3.6 Spatial structure

The classes occupied distinct geographies (Fig. 3a; Fig. 4). The chronically low class was predominantly urban (65.0% of tracts) yet close to grocery retail (mean 3.8 km from the tract population-weighted centroid to the nearest grocery store in 2024), as was the persistently-constrained class (55.3% urban; 2.6 km). The collapse classes were mostly rural and farther from grocery retail (early collapse 6.2 km, late collapse 7.9 km; Table 1). Chronically low, steady-decline and stable-adequate tracts were spatially clustered (join-count z = 4.55, 3.18 and 2.62), whereas the collapse classes were not (z = 0.55 and 1.64; P = 0.29 and 0.05); in the Getis-Ord Gi* analysis of the 2024 index, still-served towns formed significant high-adequacy clusters (49 tracts), and there were no significant low-adequacy clusters, because low adequacy was the statewide background rather than a local deviation (Additional file 1: Table S11). Fig. 4 maps the 1997 and 2024 index surfaces, their change, the 2024 dollar- and grocery-store counts, and the SVI surface.

**Figure 4.**
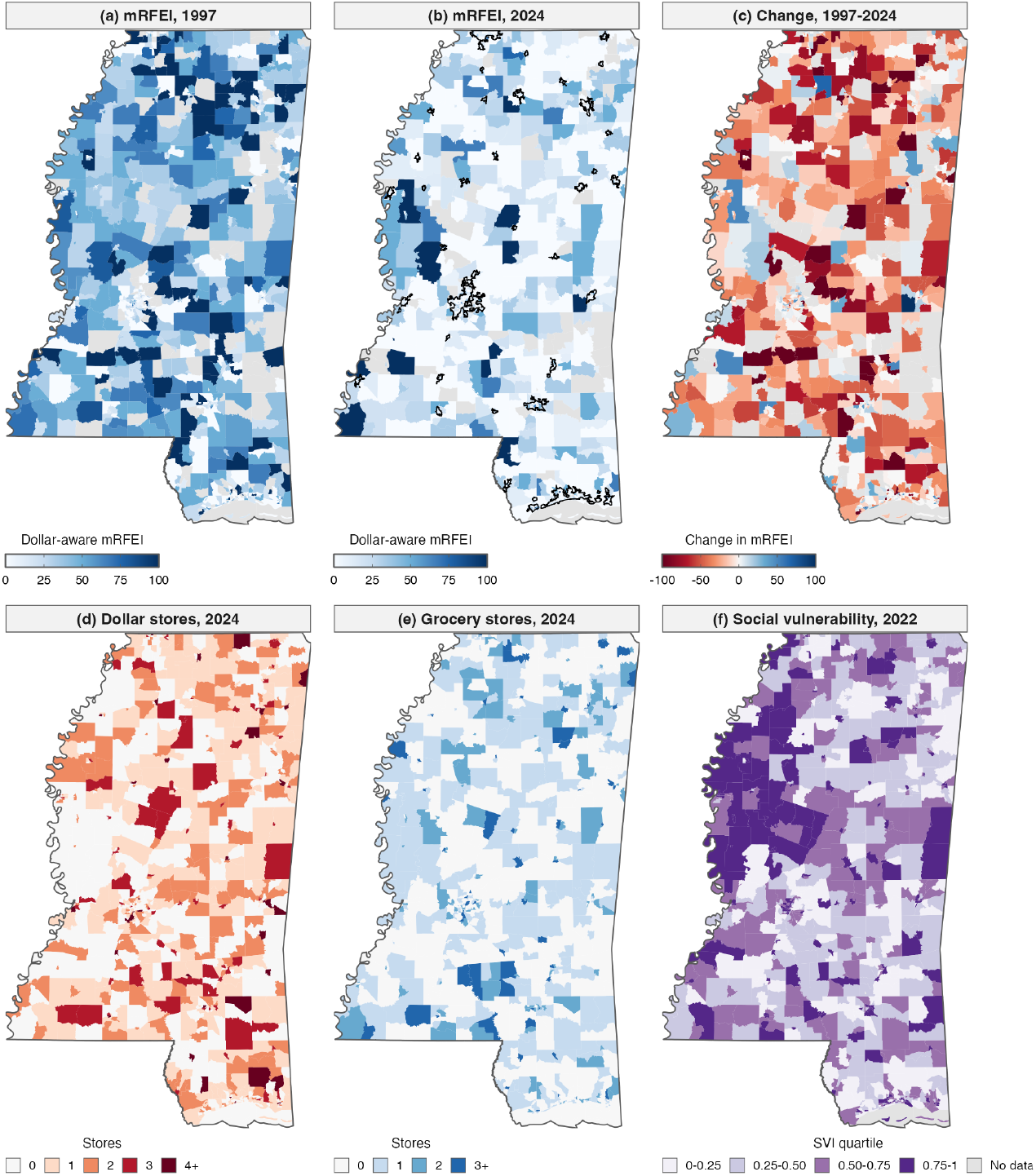
Census-tract food-retail environment in Mississippi. (a) Dollar-aware modified Retail Food Environment Index (mRFEI), 1997; (b) 2024, with 2020 Census urban areas outlined; (c) change, 1997–2024; (d) dollar stores, 2024; (e) grocery stores, 2024; (f) CDC/ATSDR Social Vulnerability Index 2022, by quartile. In (a)–(c), grey marks tracts with no food retailer (index undefined). All panels 2020 tract boundaries. Source: Data Axle; US Census Bureau TIGER/Line 2022; CDC/ATSDR SVI 2022.

## 4. Discussion

This study examined how the retail food environment of Mississippi changed over 27 years and how a single classification decision, whether dollar stores are counted, shapes the measured exposure. Three findings stand out. First, counting dollar stores barely changed how tracts ranked, but it lowered index values where dollar stores and healthy outlets coexisted, by a margin that doubled over the period, and determined whether a growing set of tracts could be scored at all. Second, a robust typology separated tracts with persistently low scores, mostly urban and close to grocery stores, from tracts whose food environments collapsed, mostly rural and far from them. Third, collapse tracts were indistinguishable from others on present-day social characteristics.

For studies that use the mRFEI as an exposure, the consequences of omitting dollar stores depend on how the index is used. Because within-year ranks were nearly unchanged, analyses that use quantiles or ranks of the index would be little affected. Analyses that use index values or fixed cut-offs would assign higher scores to tracts that contain both dollar stores and healthy outlets, by about 9 points by 2024 in such tracts. Analytic samples would also differ: a study using the standard index would drop tracts whose only retailers are dollar stores, which in Mississippi were concentrated in less vulnerable tracts, so the omission is not random with respect to characteristics that also relate to diet and weight. And because the size of the difference varied with social vulnerability, being largest in the least vulnerable tracts, the measurement difference is correlated with population characteristics rather than random. Such differences could bias associations with diet or obesity in either direction. We do not claim that either index is correct; the choice should be explicit and, where results depend on it, reported both ways.

The trajectory typology converts a long, complex panel into a small set of exposure histories suited to life-course analysis. Its most robust distinction is between tracts that were low throughout and tracts that lost an adequate food environment. The collapse classes showed no association with present-day vulnerability or urbanicity, so a cross-sectional measure would not distinguish a tract whose food environment collapsed from one that never had an adequate one, although their residents may have had very different exposure histories. The split between early and late collapse was less robust: it depended on tracts with few retailers, in which the loss of a single grocery store moves the index substantially, so we treat timing as a hypothesis for future research rather than an established feature. Moderate agreement between model specifications is common in latent class trajectory modelling [28], which is why we report the main classifications under both specifications and base our conclusions on the features that both share. Tract-level exposure histories also assume that residents stayed in place; residential mobility varies across households, so individual exposure histories will differ from those of the tracts in which people currently live.

The typology also distinguishes two forms of poor food environment. The chronically low and persistently-constrained classes were predominantly urban and close to grocery stores but scored low throughout: the configuration of a food swamp, in which less-healthy outlets dominate the retail mix rather than healthy outlets being absent [5]. The collapse classes were rural, far from grocery stores and not spatially clustered: the configuration of a food desert produced by dispersed local losses rather than a contiguous decline front. Measures based solely on distance, or on counts of a single outlet type, would conflate these environments [4,3]. Social vulnerability was a weaker and less consistent correlate than urbanicity: persistently-constrained and steady-decline membership rose modestly with 2022 vulnerability, but the association weakened with 2010 SVI and, in the components model, no single component was independently associated. Because SVI was measured at the end of the period, these cross-sectional associations could partly reflect the food-environment changes themselves. National studies report that dollar stores are concentrated in low-income and rural communities and in communities of color [10,6,7]; in this panel, the trajectory of the overall food environment was more closely tied to urbanicity than to social composition, consistent with arguments that unequal food access reflects broader retail inequality rather than the presence or absence of particular stores [52].

These patterns have implications beyond exposure measurement. Most children in classified tracts lived in food environments that declined, collapsed or stayed low over the study period. From a food-justice perspective, the long-run loss of food retail in these communities can be understood as disinvestment rather than as the outcome of individual choices [52], a view echoed by residents who describe dollar stores as meeting some needs while investing little in the communities they serve [53]. International human-rights instruments define the right to adequate food to include physical and economic access to it [54], so persistent or worsening local food environments are a question of rights as well as of public health. Programme rules shape this landscape: dollar stores are widely authorised to redeem SNAP benefits [13], whereas WIC authorisation requires minimum stocks of specified foods [19], so where dollar stores replace grocery stores, WIC families may lose redemption options faster than SNAP families, and programme rules can shape what authorised stores carry: after the 2009 revision of the WIC food packages, WIC-authorised stores in Texas expanded shelf space for fruit, vegetables and whole grains [55]. Treating all dollar stores as less healthy is also a simplification whose accuracy may vary over time and place: some dollar stores offer fresh produce of comparable quality at lower prices [18], and households that shop at dollar stores may compensate with healthier purchases elsewhere [17]. An intermediate classification, or one informed by in-store produce availability where it can be measured, is a priority for future work.

Mississippi combines high poverty, a large rural population and rapid dollar-store expansion, so the magnitudes reported here should not be assumed to hold elsewhere. Some findings are likely to generalise: that excluding a fast-growing outlet type changes index levels and measurability more than rank, that changes in vendor coverage can create artificial jumps in commercial data, that ratio indices built from few outlets are volatile, and that urban and rural tracts follow different paths. Others are more context-dependent, including the size of the index difference, the specific classes and their prevalence, and their social patterning, which will depend on local retail structure, settlement patterns and data sources. In countries without a comparable dollar-store sector, the same measurement question applies to whichever limited-assortment or discount formats are expanding.

### 4.1 Implications for food-environment measurement

These results suggest six practices for longitudinal food-environment measurement. First, state how discount and limited-assortment outlets are classified and, where results depend on it, report the index both ways. Second, report how many units each index cannot score, and which units they are. Third, before interpreting year-to-year change, check total records and new-record rates for changes in vendor coverage [44]. Fourth, avoid defining categories with fields that shift with vendor coding, such as employee size, or test their influence. Fifth, identify units with few outlets, in which a single opening or closure moves a ratio index substantially, and test robustness to excluding them. Sixth, model bounded indices with methods suited to floor and ceiling effects [46], and report trajectory models against established guidelines [51].

### 4.2 Limitations

This study has limitations. The analysis is descriptive and associational; we do not estimate the causal effect of dollar-store entry on other retailers or on any health outcome, and the associations with social vulnerability are cross-sectional. The index measures the mix of outlet types rather than the foods stocked, purchased or eaten, and classifying all dollar stores as less healthy and all grocery stores as healthy are simplifications; the index was highly sensitive to the treatment of small grocery stores, which could not be defined reliably over time. Classification was rule-based: we validated dollar-store counts against company-reported totals and corrected the misclassifications we identified in a sensitivity analysis, but no independent manual double-coding of establishments was performed, and the correctable errors we found, chiefly convenience-format stores carrying grocery codes, would bias the index towards overstating healthy access. Commercial databases also miss some small and independent outlets and date openings and closures imperfectly [21,33,22]. The trajectory classes are latent-model constructs whose boundaries are probabilistic, and the timing distinction between the collapse classes depended on tracts with few retailers. Distance to grocery stores was measured as straight-line distance to stores within Mississippi, which understates travel where road networks are sparse and ignores stores across state lines.

Most importantly, we did not measure diet, dietary intake or body weight. This paper characterises the exposure rather than its health consequences; a deteriorating or persistently poor food-retail environment is a reason to study dietary and obesity outcomes, not evidence that such outcomes have occurred. The study was conducted in a single state, so the observed magnitudes are not nationally representative, and comparable long-run establishment data remain scarce for other states.

This paper provides the exposure measures for a planned longitudinal study of childhood obesity in Mississippi, in which tract-level, time-varying food-environment trajectories will be examined in relation to children’s weight and related outcomes.

## 5. Conclusions

Between 1997 and 2024, the food-retail environment of Mississippi reorganised as grocery stores declined and dollar stores proliferated. Whether dollar stores are counted changed the level of a widely used food-environment index and, increasingly, whether a tract could be scored at all, but not how tracts ranked. A 27-year typology, robust to a range of sensitivity analyses, distinguished urban tracts with persistently low scores from rural tracts whose food environments collapsed, differences that cross-sectional measures cannot capture. These tools, and the measurement lessons that accompany them, provide a transferable, time-varying exposure framework for research on diet, food access and diet-related disease, including childhood obesity.

## Data Availability

The establishment-level business records analysed here were obtained under licence from Data Axle (formerly Infogroup) and are not publicly available; the licence does not permit redistribution of the underlying records. Equivalent historical files can be obtained from Data Axle (https://www.data-axle.com) under a commercial licence or institutional subscription. All secondary data are publicly available from their cited sources. The analysis code is available from the corresponding author on request.

## Abbreviations

ACS: American Community Survey
APPA: average posterior probability of assignment
ARI: adjusted Rand index
ATSDR: Agency for Toxic Substances and Disease Registry
BIC: Bayesian Information Criterion
CDC: Centers for Disease Control and Prevention
CI: confidence interval
GRoLTS: Guidelines for Reporting on Latent Trajectory Studies
IRB: institutional review board
mRFEI: modified Retail Food Environment Index
NAICS: North American Industry Classification System
OCC: occupancy odds ratio
RRR: relative risk ratio
SIC: Standard Industrial Classification
SNAP: Supplemental Nutrition Assistance Program
STROBE: Strengthening the Reporting of Observational Studies in Epidemiology
SVI: Social Vulnerability Index
WIC: Special Supplemental Nutrition Program for Women, Infants, and Children

## Declarations

### Ethics approval and consent to participate

This study did not involve human participants, human tissue, or identifiable personal data; therefore, institutional review board (IRB) approval was not required.

### Consent for publication

Not applicable.

### Authors’ contributions

S.V.M. and E.D.W. conceptualized the study. S.V.M. developed the methodology, wrote the software, curated the data, conducted the formal analysis and investigation, and prepared the visualizations. E.D.W. provided resources, supervised the work, administered the project, and acquired funding. S.V.M. wrote the original draft. S.V.M., S.L., and E.D.W. validated the results and reviewed and edited the manuscript. All authors read and approved the final manuscript.

## Acknowledgements

Not applicable.

## Additional files

Additional file 1 (.docx). Supplementary Methods, Tables S1–S11 and Figures S1–S4.

Additional file 2 (.docx). STROBE-nut checklist.

Additional file 3 (.docx). GRoLTS checklist for latent trajectory studies.

