## additional file 1 for "A dollar-aware food-environment index and a 27-year trajectory typology: a measurement foundation for diet and childhood-obesity research in Mississippi, 1997–2024"

 doi:10.64898/2026.08.20.26360912

### Additional file 1

#### Contents


- Supplementary Methods
- Table S1. Identification of dollar stores: signals and industry codes
- Table S2. Dollar General counts compared with company-reported Mississippi store totals, 2017–2024
- Table S3. Systematic misclassification identified in the classification review, by year
- Table S4. Statewide food-retail establishments by category, 1997–2024
- Table S5. Database coverage by year
- Table S6. Standard and dollar-aware indices compared, by year
- Table S7. Model selection, one to eight classes, spline-link and linear specifications
- Table S8. Six-class solutions: classification adequacy, model fit and class characteristics
- Table S9. Class retention in the low-count sensitivity analyses
- Table S10. Multinomial logistic regression of class membership: all specifications
- Table S11. Spatial clustering of trajectory classes and index values
- Figures S1–S4

### Supplementary Methods

#### S1. Classification of retail categories

Each annual Data Axle file was processed with the same rules. Dollar stores were identified from three signals in each record: establishment name, matched against the national and regional chains listed in the Methods and a screened list of local dollar-store chains with more than two Mississippi locations; corporate-parent identifier, which recovers chain stores whose names were recorded inconsistently; and NAICS code, which served as a confirming signal (Table S1). Records matching more than one chain (113, 0.6%) were assigned to a single chain by a fixed labelling rule; this does not affect dollar-store counts.

Grocery stores, supercentres and convenience stores (including gas stations with a store) were classified by name and industry code. Non-food establishments carrying food-retail codes (florists, pharmacies, restaurants, used-car dealers, tobacco retailers, frozen-food wholesalers and hardware stores) were excluded from the grocery category, while discount-grocery formats were retained. Large-format supercentres generated several records at one location in later years; only the principal record per location was retained, which reduced supercentre establishment-years from 2,033 to 1,665. Discontinuities in grocery and convenience counts around the 2008 SIC-to-NAICS transition reflected re-coding rather than retail change and were resolved with exact NAICS sub-codes and screening on descriptive industry text and establishment name. Fast-food outlets were limited-service restaurants identified by name and industry code, excluding outlets located inside other stores. Across 164,326 establishment-years in the five index categories, one establishment-year was counted in two categories (grocery and convenience).

#### S2. Index definitions

For tract i in year t, with H healthy retailers (grocery stores and supercentres), L less-healthy retailers (convenience stores and fast-food outlets) and D dollar stores, the standard index is mRFEI $= 100 \times H / (H + L)$ and the dollar-aware index is $100 \times H / (H + L + D)$. Each index is undefined when its denominator is zero. The difference between them is zero when $D = 0$ and also when $H = 0$, so it can be non-zero only in tract-years containing both a dollar store and a healthy outlet. Tract-years in which $D > 0$ and $H + L = 0$ are scored 0 by the dollar-aware index and are undefined under the standard index. The size-based sensitivity definition moves grocery stores with three or fewer employees from H to L, following the CDC definition.

#### S3. Trajectory models

Latent class mixed models were fitted with the lcmm package in R to the tract-level dollar-aware index, with time scaled as (year − 1997)/10, cubic class-specific fixed effects and no within-class random effects. The primary specification used lcmm() with a monotone spline link (five equidistant knots, "5-equi-splines"); the linear specification used hlme(). Models with one to eight classes were each fitted from 24 random starts (gridsearch), and the best start was iterated to convergence. Tracts were assigned to the class with the highest posterior probability. Adequacy was assessed by relative entropy, the class-specific average posterior probability of assignment (APPA) and the occupancy odds ratio (OCC), with APPA $\geq 0.7$ and OCC $\geq 5$ regarded as adequate. Agreement between classifications was summarised by the adjusted Rand index (ARI) and by the percentage of tracts retaining their class after each class in the comparison solution was matched to the main-solution class with which it shared most tracts. Sensitivity solutions were fitted from new random starts or, where stated, from the main solution's estimates.

##### S4. Social vulnerability and urbanicity

The 2022 CDC/ATSDR Social Vulnerability Index (overall percentile, Mississippi state ranking) was used for 2020 tracts. For the earlier-period sensitivity analysis, SVI 2010 (Mississippi state ranking, 2010 tracts) was assigned to 2020 tracts with the Census Bureau 2020–2010 tract relationship file, weighting each contributing 2010 tract by its share of the 2020 tract's land area; 872 of 878 tracts received a value (median dominant-part share 1.00; 279 tracts were built from more than one 2010 tract). The two measures correlated at 0.71 (Pearson and Spearman). A tract was urban if its 2020 population-weighted centroid fell within a 2020 Census urban area. Multinomial models used stable adequate as the reference class; 95% confidence intervals were obtained from 1,000 resamples of Mississippi's 82 counties with replacement (percentile intervals), alongside model-based Wald intervals.

##### S5. Spatial statistics and distances

Join-count statistics for each class indicator used first-order queen contiguity on 2020 tract boundaries and tested whether same-class tracts adjoin more often than expected under spatial randomness. The Getis-Ord Gi* statistic was computed on the 2024 dollar-aware index with binary queen-contiguity weights including each tract itself; $|z| \geq 1.96$ and $\geq 2.576$ identified clusters at the 95% and 99% levels. Distances are straight-line distances from each tract's 2020 population-weighted centroid to the nearest operating grocery store, and to the nearest grocery store or supercentre, in Mississippi in 2024.

#### Supplementary Tables

**Table S1.** Identification of dollar stores: signals and industry codes.

| Item | Detail | Records (%) |
|---|---|---|
| Name and corporate parent | Both signals matched | 14,176 (76.3) |
| Name only | Chain name matched; parent identifier absent or different | 3,436 (18.5) |
| Corporate parent only | Parent identifier matched; name recorded inconsistently (e.g. Fred's, 2004–2012) | 970 (5.2) |
| **All dollar-store establishment-years** | | **18,582 (100.0)** |
| Confirmed by NAICS code | Variety- or general-merchandise NAICS code present | 18,254 (98.2) |
| NAICS code alone (not used for inclusion) | Records with the same codes but no chain match; chiefly supercentres and department and apparel stores | 14,284 |
| Industry codes, SIC | Dollar General: 531102 (department stores) to 2004, 531104 in 2005, 533101 (variety stores) by 2016; Family Dollar: 531102 in 1997–2003 | |
| Industry codes, NAICS | Prefixes 452990 and 45211 (earlier vintages); 452319 and 45299013 (2017 vintage); 455219 and 45521903 (2022 vintage); 45521939 (Ollie's Bargain Outlet) | |
| Chains | Dollar General (incl. DG Market, pOpshelf), Family Dollar, Dollar Tree, Fred's (discount stores), Big Lots, Five Below, Ollie's Bargain Outlet, Super 10, and local dollar-store chains with more than two Mississippi locations; Dirt Cheap excluded | |

*Note:* Establishment-years are dollar-store records in the 28 annual files, 1997–2024. Data Axle assigns six-digit SIC and eight-digit NAICS extensions. Contemporary dollar-store NAICS codes alone would have missed 77–98% of dollar stores before 2006 and about 3–6% by the early 2020s. Excluding Big Lots and Ollie's Bargain Outlet reduces dollar-store establishment-years by 2.3%. Source: Data Axle.

**Table S2.** Dollar General counts compared with company-reported Mississippi store totals, 2017–2024.

| Year | This study | Company-reported | Difference | Difference, % |
|---|---|---|---|---|
| 2017 | 463 | 483 | −20 | −4.1 |
| 2018 | 473 | 512 | −39 | −7.6 |
| 2019 | 518 | 538 | −20 | −3.7 |
| 2020 | 543 | 563 | −20 | −3.6 |
| 2021 | 550 | 587 | −37 | −6.3 |
| 2022 | 582 | 621 | −39 | −6.3 |
| 2023 | 652 | 657 | −5 | −0.8 |
| 2024 | 637 | 668 | −31 | −4.6 |

*Note:* Company-reported counts are Mississippi store totals from Dollar General Form 10-K filings (Item 2, Properties), reported as of early March of the following year and aligned to the preceding calendar year. Differences are relative to the company figure (mean absolute difference 4.6%): a slight, consistent undercount with no evidence of overcounting. Source: Data Axle; Dollar General Corporation.

**Table S3.** Systematic misclassification identified in the classification review, by year.

| Year | Convenience records | EV charging stations | Grocery records | Convenience-format stores | Supercentre records | Neighborhood Markets | Café listings |
|---|---|---|---|---|---|---|---|
| 1997 | 2,313 | 0 | 1,616 | 41 | 12 | 0 | 0 |
| 1998 | 2,185 | 0 | 1,548 | 60 | 13 | 0 | 0 |
| 1999 | 2,206 | 0 | 1,525 | 66 | 18 | 0 | 0 |
| 2000 | 2,203 | 0 | 1,487 | 58 | 21 | 0 | 0 |
| 2001 | 2,356 | 0 | 1,435 | 53 | 26 | 0 | 0 |
| 2002 | 2,425 | 0 | 1,421 | 48 | 33 | 0 | 0 |
| 2003 | 2,554 | 0 | 1,383 | 27 | 27 | 0 | 0 |
| 2004 | 2,590 | 0 | 1,384 | 29 | 48 | 0 | 0 |
| 2005 | 2,585 | 0 | 1,355 | 34 | 53 | 0 | 0 |
| 2006 | 2,491 | 0 | 1,240 | 23 | 50 | 0 | 0 |
| 2007 | 2,464 | 0 | 1,196 | 27 | 52 | 0 | 0 |
| 2008 | 2,475 | 0 | 1,105 | 19 | 51 | 0 | 0 |
| 2009 | 2,474 | 0 | 1,076 | 26 | 60 | 1 | 0 |
| 2010 | 2,406 | 0 | 1,056 | 24 | 68 | 1 | 0 |
| 2011 | 2,382 | 1 | 974 | 19 | 66 | 1 | 0 |
| 2012 | 2,504 | 1 | 1,105 | 25 | 69 | 1 | 0 |
| 2013 | 2,491 | 15 | 1,140 | 28 | 70 | 1 | 0 |
| 2014 | 2,527 | 21 | 1,093 | 27 | 74 | 1 | 0 |
| 2015 | 2,519 | 25 | 1,062 | 31 | 85 | 12 | 0 |
| 2016 | 2,524 | 28 | 951 | 23 | 79 | 6 | 0 |
| 2017 | 2,448 | 34 | 877 | 21 | 81 | 9 | 0 |
| 2018 | 2,402 | 37 | 871 | 41 | 83 | 10 | 0 |
| 2019 | 2,324 | 50 | 830 | 47 | 85 | 10 | 0 |
| 2020 | 2,167 | 55 | 775 | 46 | 86 | 10 | 0 |
| 2021 | 2,230 | 89 | 756 | 43 | 86 | 10 | 0 |
| 2022 | 2,260 | 109 | 760 | 47 | 86 | 10 | 0 |
| 2023 | 2,205 | 93 | 740 | 46 | 92 | 10 | 7 |
| 2024 | 2,179 | 94 | 716 | 45 | 91 | 10 | 7 |
| **Total** | **66,889** | **652** | **31,477** | **1,024** | **1,665** | **103** | **14** |

*Note:* EV charging stations carried gas-station codes; convenience-format stores (e.g. Keith's Superstore, E-Z Serve, Mac's Mini-Mart, "One Stop" stores) carried vendor grocery codes; Walmart Neighborhood Markets (a grocery format) were classified as supercentres, which leaves the index unchanged because both categories count as healthy; café listings within warehouse clubs were classified as supercentres. Correcting all four changed 5.69% of tract-year index values (mean change −0.74 points; Spearman correlation with the uncorrected index 0.978); class agreement with the main solutions was ARI 0.88 (spline link; 94.5% of tracts retained their class) and 0.84 (linear; 93.5%). Source: Data Axle.

**Table S4.** Statewide food-retail establishments by category, 1997–2024.

| Year | Grocery | Supercentre | Convenience | Dollar | Fast food | Total | Grocery, % | Dollar, % | Dollar:grocery |
|---|---|---|---|---|---|---|---|---|---|
| 1997 | 1,616 | 12 | 2,313 | 315 | 1,355 | 5,611 | 28.8 | 5.6 | 0.19 |
| 1998 | 1,548 | 13 | 2,185 | 334 | 1,366 | 5,446 | 28.4 | 6.1 | 0.22 |
| 1999 | 1,525 | 18 | 2,206 | 381 | 1,387 | 5,517 | 27.6 | 6.9 | 0.25 |
| 2000 | 1,487 | 21 | 2,203 | 372 | 1,369 | 5,452 | 27.3 | 6.8 | 0.25 |
| 2001 | 1,435 | 26 | 2,356 | 442 | 1,502 | 5,761 | 24.9 | 7.7 | 0.31 |
| 2002 | 1,421 | 33 | 2,425 | 462 | 1,516 | 5,857 | 24.3 | 7.9 | 0.33 |
| 2003 | 1,383 | 27 | 2,554 | 499 | 1,402 | 5,865 | 23.6 | 8.5 | 0.36 |
| 2004 | 1,384 | 48 | 2,590 | 527 | 1,380 | 5,929 | 23.3 | 8.9 | 0.38 |
| 2005 | 1,355 | 53 | 2,585 | 546 | 1,458 | 5,997 | 22.6 | 9.1 | 0.4 |
| 2006 | 1,240 | 50 | 2,491 | 544 | 1,470 | 5,795 | 21.4 | 9.4 | 0.44 |
| 2007 | 1,196 | 52 | 2,464 | 571 | 1,488 | 5,771 | 20.7 | 9.9 | 0.48 |
| 2008 | 1,105 | 51 | 2,475 | 588 | 1,514 | 5,733 | 19.3 | 10.3 | 0.53 |
| 2009 | 1,076 | 60 | 2,474 | 595 | 1,542 | 5,747 | 18.7 | 10.4 | 0.55 |
| 2010 | 1,056 | 68 | 2,406 | 620 | 1,542 | 5,692 | 18.6 | 10.9 | 0.59 |
| 2011 | 974 | 66 | 2,382 | 624 | 1,525 | 5,571 | 17.5 | 11.2 | 0.64 |
| 2012 | 1,105 | 69 | 2,504 | 708 | 1,646 | 6,032 | 18.3 | 11.7 | 0.64 |
| 2013 | 1,140 | 70 | 2,491 | 745 | 1,728 | 6,174 | 18.5 | 12.1 | 0.65 |
| 2014 | 1,093 | 74 | 2,527 | 771 | 1,791 | 6,256 | 17.5 | 12.3 | 0.71 |
| 2015 | 1,062 | 85 | 2,519 | 802 | 1,815 | 6,283 | 16.9 | 12.8 | 0.76 |
| 2016 | 951 | 79 | 2,524 | 843 | 1,829 | 6,226 | 15.3 | 13.5 | 0.89 |
| 2017 | 877 | 81 | 2,448 | 874 | 1,801 | 6,081 | 14.4 | 14.4 | 1 |
| 2018 | 871 | 83 | 2,402 | 891 | 1,840 | 6,087 | 14.3 | 14.6 | 1.02 |
| 2019 | 830 | 85 | 2,324 | 898 | 1,886 | 6,023 | 13.8 | 14.9 | 1.08 |
| 2020 | 775 | 86 | 2,167 | 843 | 1,875 | 5,746 | 13.5 | 14.7 | 1.09 |
| 2021 | 756 | 86 | 2,230 | 861 | 1,850 | 5,783 | 13.1 | 14.9 | 1.14 |
| 2022 | 760 | 86 | 2,260 | 914 | 1,882 | 5,902 | 12.9 | 15.5 | 1.2 |
| 2023 | 740 | 92 | 2,205 | 1,007 | 1,952 | 5,996 | 12.3 | 16.8 | 1.36 |
| 2024 | 716 | 91 | 2,180 | 1,005 | 2,002 | 5,994 | 11.9 | 16.8 | 1.4 |
| **Change 1997–2024, %** | **−55.7** | **+658.3** | **−5.8** | **+219.0** | **+47.7** | **+6.8** | | | |
| Annual growth, % | −3.0 | +6.9 | −0.1 | +4.1 | +1.5 | +0.3 | | | |

*Note:* Counts are operating establishments in each annual file, statewide (including one 2003 grocery store without coordinates that cannot be assigned to a tract). Percentages are shares of the five categories combined. Annual growth is the compound rate from a regression of log counts on year. Dollar stores first outnumbered grocery stores in 2018. Source: Data Axle.

**Table S5.** Database coverage by year.

| Year | All records | Change, % | Grocery, change % | Convenience, change % | Dollar, change % | Fast food, change % | Grocery records new to database, % | Grocery per 10,000 records |
|---|---|---|---|---|---|---|---|---|
| 1997 | 97,680 | | | | | | | 165.4 |
| 1998 | 97,841 | +0.2 | −4.2 | −5.5 | +6.0 | +0.8 | 6.1 | 158.2 |
| 1999 | 100,366 | +2.6 | −1.5 | +1.0 | +14.1 | +1.5 | 6.8 | 151.9 |
| 2000 | 100,196 | −0.2 | −2.5 | −0.1 | −2.4 | −1.3 | 1.3 | 148.4 |
| 2001 | 117,579 | +17.3 | −3.5 | +6.9 | +18.8 | +9.7 | 14.4 | 122 |
| 2002 | 117,319 | −0.2 | −1.0 | +2.9 | +4.5 | +0.9 | 8.4 | 121.1 |
| 2003 | 116,802 | −0.4 | −2.7 | +5.3 | +8.0 | −7.5 | 7.4 | 118.4 |
| 2004 | 117,062 | +0.2 | +0.1 | +1.4 | +5.6 | −1.6 | 9.8 | 118.2 |
| 2005 | 119,361 | +2.0 | −2.1 | −0.2 | +3.6 | +5.7 | 9.5 | 113.5 |
| 2006 | 119,543 | +0.2 | −8.5 | −3.6 | −0.4 | +0.8 | 10.6 | 103.7 |
| 2007 | 120,057 | +0.4 | −3.5 | −1.1 | +5.0 | +1.2 | 10.7 | 99.6 |
| 2008 | 119,510 | −0.5 | −7.6 | +0.4 | +3.0 | +1.7 | 10.4 | 92.5 |
| 2009 | 118,748 | −0.6 | −2.6 | +0.0 | +1.2 | +1.8 | 10 | 90.6 |
| 2010 | 119,566 | +0.7 | −1.9 | −2.7 | +4.2 | +0.0 | 11.5 | 88.3 |
| 2011 | 116,155 | −2.9 | −7.8 | −1.0 | +0.6 | −1.1 | 8.5 | 83.9 |
| 2012 | 126,610 | +9.0 | +13.4 | +5.1 | +13.5 | +7.9 | 18.4 | 87.3 |
| 2013 | 137,002 | +8.2 | +3.2 | −0.5 | +5.2 | +5.0 | 12 | 83.2 |
| 2014 | 139,437 | +1.8 | −4.1 | +1.4 | +3.5 | +3.6 | 7.5 | 78.4 |
| 2015 | 141,100 | +1.2 | −2.8 | −0.3 | +4.0 | +1.3 | 9.4 | 75.3 |
| 2016 | 137,153 | −2.8 | −10.5 | +0.2 | +5.1 | +0.8 | 6.2 | 69.3 |
| 2017 | 127,988 | −6.7 | −7.8 | −3.0 | +3.7 | −1.5 | 4 | 68.5 |
| 2018 | 124,045 | −3.1 | −0.7 | −1.9 | +1.9 | +2.2 | 10.7 | 70.2 |
| 2019 | 122,150 | −1.5 | −4.7 | −3.2 | +0.8 | +2.5 | 6 | 67.9 |
| 2020 | 120,687 | −1.2 | −6.6 | −6.8 | −6.1 | −0.6 | 6.3 | 64.2 |
| 2021 | 122,934 | +1.9 | −2.5 | +2.9 | +2.1 | −1.3 | 4.9 | 61.5 |
| 2022 | 128,199 | +4.3 | +0.5 | +1.3 | +6.2 | +1.7 | 5.9 | 59.3 |
| 2023 | 131,407 | +2.5 | −2.6 | −2.4 | +10.2 | +3.7 | 4.3 | 56.3 |
| 2024 | 137,662 | +4.8 | −3.2 | −1.1 | −0.2 | +2.6 | 2.2 | 52 |

*Note:* All records are every Mississippi establishment in the annual file. Grocery records new to the database have an establishment identifier not present in any earlier file. Of the 203 grocery records new in 2012, 77.3% were still listed as grocery stores in 2013, 62.6% in 2014, 54.7% in 2015 and 41.4% in 2016. Source: Data Axle.

**Table S6.** Standard and dollar-aware indices compared, by year.

| Year | Tracts scored by both | With a dollar store, % | Mean difference, all scored | Tract-years where a difference is possible | Mean difference where possible | Median where possible | Dollar-store tracts with no healthy outlet, % | Dollar-store-only tracts | Spearman | Moved >10 percentile points, % |
|---|---|---|---|---|---|---|---|---|---|---|
| 1997 | 756 | 27.4 | 1.12 | 188 | 4.52 | 3.12 | 9.2 | 1 | 0.995 | 2.2 |
| 1998 | 762 | 28.3 | 1.16 | 195 | 4.52 | 2.87 | 9.7 | 2 | 0.996 | 1.4 |
| 1999 | 775 | 29.8 | 1.33 | 209 | 4.93 | 3.1 | 9.5 | 2 | 0.995 | 1.3 |
| 2000 | 773 | 29.2 | 1.32 | 204 | 5.01 | 3.03 | 9.7 | 1 | 0.995 | 1.4 |
| 2001 | 790 | 32.8 | 1.55 | 229 | 5.35 | 3.16 | 11.6 | 2 | 0.993 | 2 |
| 2002 | 793 | 34.9 | 1.63 | 249 | 5.19 | 3.27 | 10.1 | 1 | 0.994 | 1.4 |
| 2003 | 801 | 37.3 | 1.64 | 260 | 5.06 | 3.33 | 13 | 0 | 0.993 | 1.2 |
| 2004 | 809 | 38.3 | 1.85 | 279 | 5.37 | 3.53 | 10 | 0 | 0.992 | 1.5 |
| 2005 | 802 | 38.8 | 1.91 | 277 | 5.54 | 3.33 | 10.9 | 3 | 0.992 | 1.7 |
| 2006 | 797 | 40.4 | 1.81 | 284 | 5.08 | 3.33 | 11.8 | 3 | 0.994 | 2 |
| 2007 | 795 | 42.6 | 1.82 | 281 | 5.16 | 3.33 | 17.1 | 5 | 0.993 | 2.4 |
| 2008 | 796 | 44.1 | 1.8 | 287 | 4.98 | 3.33 | 18.2 | 7 | 0.994 | 2.3 |
| 2009 | 797 | 44.8 | 1.9 | 292 | 5.19 | 3.33 | 18.2 | 5 | 0.993 | 2.4 |
| 2010 | 800 | 46.5 | 2.01 | 297 | 5.42 | 3.33 | 20.2 | 4 | 0.994 | 1.5 |
| 2011 | 795 | 47.9 | 2.13 | 293 | 5.77 | 3.33 | 23.1 | 4 | 0.995 | 0.5 |
| 2012 | 805 | 52 | 2.46 | 332 | 5.95 | 3.57 | 20.8 | 4 | 0.993 | 2.1 |
| 2013 | 802 | 52.6 | 2.9 | 337 | 6.91 | 3.9 | 20.1 | 7 | 0.991 | 3.4 |
| 2014 | 805 | 53.3 | 2.63 | 337 | 6.29 | 3.57 | 21.4 | 9 | 0.993 | 0.6 |
| 2015 | 806 | 56.2 | 2.86 | 346 | 6.65 | 3.55 | 23.6 | 8 | 0.993 | 1.6 |
| 2016 | 794 | 59.2 | 2.86 | 342 | 6.64 | 3.33 | 27.2 | 16 | 0.994 | 0.9 |
| 2017 | 782 | 62.5 | 3.3 | 357 | 7.23 | 3.57 | 27 | 23 | 0.993 | 1.8 |
| 2018 | 782 | 63.8 | 3.48 | 365 | 7.45 | 4.04 | 26.9 | 26 | 0.993 | 1.5 |
| 2019 | 780 | 65.4 | 4.16 | 362 | 8.96 | 4.17 | 29 | 28 | 0.993 | 1.8 |
| 2020 | 774 | 65.6 | 3.67 | 339 | 8.38 | 3.9 | 33.3 | 26 | 0.994 | 1.7 |
| 2021 | 777 | 67.1 | 3.73 | 346 | 8.38 | 4.17 | 33.6 | 25 | 0.994 | 1.3 |
| 2022 | 785 | 68 | 3.87 | 353 | 8.61 | 4.55 | 33.9 | 27 | 0.994 | 1.1 |
| 2023 | 773 | 69.3 | 3.99 | 344 | 8.96 | 4.29 | 35.8 | 31 | 0.993 | 1.3 |
| 2024 | 774 | 69.9 | 4.07 | 348 | 9.05 | 4.29 | 35.7 | 32 | 0.993 | 2.5 |

*Note:* Differences are standard minus dollar-aware index (0–100 points). A difference is possible only in tract-years with at least one dollar store and at least one healthy outlet. Dollar-store-only tracts have dollar stores as their only food retailers: scored 0 by the dollar-aware index and undefined by the standard index (302 tract-years in 72 tracts, 1997–2024). Rank statistics are computed within each year among tracts both indices score. Source: Data Axle.

**Table S7.** Model selection, one to eight classes, spline-link and linear specifications.

| Model | K | Parameters | Log-likelihood | BIC | SABIC | BIC change, % | Relative entropy | Min. APPA | Min. OCC | Smallest class, n (%) |
|---|---|---|---|---|---|---|---|---|---|---|
| Spline link | 1 | 10 | −95,180.2 | 190,427.5 | 190,395.7 | | | 1.000 | | 817 (100) |
| | 2 | 15 | −91,588.8 | 183,278.2 | 183,230.6 | −3.75 | 0.937 | 0.982 | 47.2 | 374 (45.8) |
| | 3 | 20 | −89,740.4 | 179,614.9 | 179,551.4 | −2.00 | 0.965 | 0.978 | 56.9 | 174 (21.3) |
| | 4 | 25 | −88,368.0 | 176,903.6 | 176,824.2 | −1.51 | 0.968 | 0.973 | 56.3 | 139 (17) |
| | 5 | 30 | −87,807.0 | 175,815.2 | 175,719.9 | −0.61 | 0.966 | 0.962 | 105.3 | 76 (9.3) |
| | 6 | 35 | −87,421.2 | 175,077.1 | 174,966.0 | −0.42 | 0.964 | 0.953 | 80.7 | 64 (7.8) |
| | 7 | 40 | −87,068.3 | 174,404.8 | 174,277.8 | −0.38 | 0.962 | 0.954 | 93.7 | 55 (6.7) |
| | 8 | 45 | −86,779.5 | 173,860.8 | 173,717.9 | −0.31 | 0.958 | 0.939 | 68.2 | 53 (6.5) |
| Linear | 1 | 5 | −102,437.3 | 204,908.2 | 204,892.3 | | | 1.000 | | 817 (100) |
| | 2 | 10 | −98,154.3 | 196,375.6 | 196,343.9 | −4.16 | 0.978 | 0.989 | 71.4 | 215 (26.3) |
| | 3 | 15 | −96,533.7 | 193,168.0 | 193,120.4 | −1.63 | 0.967 | 0.970 | 63.2 | 94 (11.5) |
| | 4 | 20 | −95,670.8 | 191,475.6 | 191,412.1 | −0.88 | 0.967 | 0.969 | 57.5 | 63 (7.7) |
| | 5 | 25 | −94,970.9 | 190,109.5 | 190,030.1 | −0.71 | 0.944 | 0.947 | 33.9 | 46 (5.6) |
| | 6 | 30 | −94,472.0 | 189,145.1 | 189,049.8 | −0.51 | 0.948 | 0.942 | 31.2 | 46 (5.6) |
| | 7 | 35 | −94,148.2 | 188,531.1 | 188,419.9 | −0.33 | 0.950 | 0.940 | 31.5 | 29 (3.5) |
| | 8 | 40 | −93,874.4 | 188,017.1 | 187,890.1 | −0.27 | 0.951 | 0.944 | 35.4 | 23 (2.8) |

*Note:* Latent class mixed models of the dollar-aware index, 817 tracts, 22,025 tract-years, each fitted from 24 random starts. K, number of classes. BIC change is relative to the solution with one fewer class. APPA, average posterior probability of assignment; OCC, occupancy odds ratio; SABIC, sample-size-adjusted BIC. The six-class solution is the smallest that separates early from late collapse; the seven-class spline-link solution split 55 tracts from the persistently-constrained (44) and chronically low (11) classes while retaining 93–98% of each other class. Source: Data Axle.

**Table S8.** Six-class solutions: classification adequacy, model fit and class characteristics.

| Model | Class | Tracts, n (%) | APPA | OCC | Mean absolute error | Index 1997, observed (predicted) | Index 2024, observed (predicted) | Mean SVI | Urban, % | Grocery, km |
|---|---|---|---|---|---|---|---|---|---|---|
| Spline link | Stable adequate | 64 (7.8) | 0.982 | 633.8 | 2.00 | 67.6 (64.8) | 56.8 (57.2) | 0.42 | 18.8 | 5 |
| | Steady decline | 125 (15.3) | 0.971 | 186.5 | 1.36 | 47.9 (48.9) | 29.1 (27) | 0.57 | 28.8 | 4.4 |
| | Early collapse | 105 (12.9) | 0.953 | 137.1 | 3.08 | 47.4 (55.4) | 4.3 (5.7) | 0.49 | 32.4 | 6.2 |
| | Late collapse | 79 (9.7) | 0.974 | 355.8 | 3.39 | 64.7 (60.5) | 2.1 (3) | 0.46 | 21.5 | 7.9 |
| | Persistently constrained | 284 (34.8) | 0.977 | 80.7 | 0.96 | 23.7 (21.6) | 14.8 (15) | 0.58 | 55.3 | 2.6 |
| | Chronically low | 160 (19.6) | 0.983 | 231.2 | 0.93 | 9.4 (6.8) | 3.3 (4.3) | 0.42 | 65 | 3.8 |
| Linear | Stable adequate | 46 (5.6) | 0.998 | 9652.3 | 2.50 | 71 (68.9) | 61.6 (60.9) | 0.38 | 17.4 | 4.9 |
| | Steady decline | 107 (13.1) | 0.979 | 305.4 | 1.37 | 49.1 (51.7) | 33.7 (33.5) | 0.55 | 28 | 4.8 |
| | Early collapse | 91 (11.1) | 0.968 | 244.2 | 3.52 | 64.3 (75.9) | 6.5 (8.9) | 0.47 | 20.9 | 7.9 |
| | Late collapse | 55 (6.7) | 0.988 | 1179.3 | 2.12 | 77.2 (73.6) | 7.9 (9.2) | 0.4 | 23.6 | 6.3 |
| | Persistently constrained | 279 (34.1) | 0.942 | 31.2 | 0.69 | 25.2 (24.8) | 14.5 (14.3) | 0.59 | 46.2 | 3.1 |
| | Chronically low | 239 (29.3) | 0.963 | 63.3 | 0.50 | 13.3 (13.3) | 5.1 (4.9) | 0.47 | 67.4 | 3.5 |

*Note:* Relative entropy was 0.964 (spline link) and 0.948 (linear). Mean absolute error is the mean absolute difference between observed and predicted class means across years. For the spline-link model the predicted trajectories of the early- and late-collapse classes fell below 20 by 2005 and 2013; the largest single-year deviation was for the early-collapse class in 2002 (observed 44.3, predicted 29.3). SVI, CDC/ATSDR Social Vulnerability Index 2022; urban, population-weighted centroid in a 2020 Census urban area; grocery, straight-line distance to the nearest grocery store in 2024. Source: Data Axle; CDC/ATSDR SVI 2022; US Census Bureau.

**Table S9.** Class retention in the low-count sensitivity analyses.

| Class | Spline link: tracts averaging <3 retailers excluded | Spline link: tract-years with <3 retailers missing | Linear: tracts averaging <3 retailers excluded | Linear: tract-years with <3 retailers missing |
|---|---|---|---|---|
| Stable adequate | 100 (30/30) | 88.9 (32/36) | 81.2 (13/16) | 85.7 (18/21) |
| Steady decline | 83.3 (90/108) | 71.4 (80/112) | 94.2 (81/86) | 60 (54/90) |
| Early collapse | 0 (0/56) | 57.4 (35/61) | 96.2 (50/52) | 0 (0/55) |
| Late collapse | 92.9 (39/42) | 0 (0/48) | 0 (0/21) | 88 (22/25) |
| Persistently constrained | 94.7 (249/263) | 91 (243/267) | 87.9 (218/248) | 93 (238/256) |
| Chronically low | 83.3 (80/96) | 84.5 (87/103) | 100 (172/172) | 92.8 (167/180) |
| **Adjusted Rand index (tracts retained, %)** | **0.53 (82.0)** | **0.43 (76.1)** | **0.81 (89.7)** | **0.58 (79.6)** |

*Note:* Values are the percentage of each main-solution class retaining its label (tracts retained/tracts in the analysis sample) after matching classes by majority overlap. Excluding tracts averaging fewer than three food retailers per year left 595 tracts; treating tract-years with fewer than three retailers as missing and retaining tracts with at least 15 such years left 627. A 0% entry means that class merged with another class. Source: Data Axle.

| Model | Specification | Class | Term | RRR | Model-based 95% CI | Cluster-bootstrap 95% CI |
|---|---|---|---|---|---|---|

**Table S10.** Multinomial logistic regression of class membership: all specifications.

| Model | Specification | Class | Term | RRR | Model-based 95% CI | Cluster-bootstrap 95% CI |
|---|---|---|---|---|---|---|
| Spline link | SVI 2022 and urbanicity (n = 816) | Steady decline | SVI 2022, per 0.1 | 1.20 | 1.07–1.35 | 1.08–1.33 |
| | | Early collapse | SVI 2022, per 0.1 | 1.08 | 0.96–1.21 | 0.96–1.21 |
| | | Late collapse | SVI 2022, per 0.1 | 1.06 | 0.93–1.19 | 0.93–1.19 |
| | | Persistently constrained | SVI 2022, per 0.1 | 1.17 | 1.06–1.30 | 1.06–1.31 |
| | | Chronically low | SVI 2022, per 0.1 | 0.95 | 0.85–1.06 | 0.85–1.07 |
| | | Steady decline | Urban vs rural | 1.42 | 0.67–3.01 | 0.72–4.65 |
| | | Early collapse | Urban vs rural | 1.92 | 0.90–4.09 | 0.92–5.28 |
| | | Late collapse | Urban vs rural | 1.12 | 0.49–2.59 | 0.38–4.59 |
| | | Persistently constrained | Urban vs rural | 4.49 | 2.27–8.85 | 2.18–14.79 |
| | | Chronically low | Urban vs rural | 8.33 | 4.07–17.03 | 4.34–26.17 |
| Spline link | SVI components and urbanicity (n = 807) | Steady decline | Black, per 10 percentage points | 1.03 | 0.89–1.19 | 0.89–1.19 |
| | | Early collapse | Black, per 10 percentage points | 1.03 | 0.89–1.19 | 0.87–1.20 |
| | | Late collapse | Black, per 10 percentage points | 1.02 | 0.87–1.19 | 0.84–1.22 |
| | | Persistently constrained | Black, per 10 percentage points | 0.99 | 0.87–1.12 | 0.86–1.12 |
| | | Chronically low | Black, per 10 percentage points | 0.96 | 0.83–1.10 | 0.75–1.13 |
| | | Steady decline | Poverty, per 10 percentage points | 1.07 | 0.72–1.60 | 0.72–1.65 |
| | | Early collapse | Poverty, per 10 percentage points | 0.91 | 0.60–1.39 | 0.59–1.35 |
| | | Late collapse | Poverty, per 10 percentage points | 1.16 | 0.75–1.80 | 0.67–1.90 |
| | | Persistently constrained | Poverty, per 10 percentage points | 1.06 | 0.73–1.53 | 0.74–1.55 |
| | | Chronically low | Poverty, per 10 percentage points | 1.07 | 0.71–1.60 | 0.75–1.64 |
| | | Steady decline | Median income, per US$10,000 | 0.83 | 0.66–1.05 | 0.65–1.02 |
| | | Early collapse | Median income, per US$10,000 | 0.95 | 0.76–1.18 | 0.76–1.17 |
| | | Late collapse | Median income, per US$10,000 | 1.05 | 0.83–1.32 | 0.86–1.26 |
| | | Persistently constrained | Median income, per US$10,000 | 0.95 | 0.78–1.16 | 0.78–1.16 |
| | | Chronically low | Median income, per US$10,000 | 1.12 | 0.92–1.37 | 0.89–1.50 |
| | | Steady decline | Urban vs rural | 1.37 | 0.63–2.99 | 0.64–4.23 |
| | | Early collapse | Urban vs rural | 2.01 | 0.91–4.41 | 0.99–5.69 |
| | | Late collapse | Urban vs rural | 1.08 | 0.45–2.57 | 0.40–4.36 |
| | | Persistently constrained | Urban vs rural | 5.08 | 2.52–10.27 | 2.49–18.75 |

| Model | Specification | Class | Term | RRR | Model-based 95% CI | Cluster-bootstrap 95% CI |
|---|---|---|---|---|---|---|
| | | Chronically low | Urban vs rural | 8.41 | 3.99–17.71 | 4.18–28.36 |
| Spline link | SVI 2010 and urbanicity (n = 817) | Steady decline | SVI 2010, per 0.1 | 1.10 | 0.99–1.23 | 0.97–1.24 |
| | | Early collapse | SVI 2010, per 0.1 | 0.99 | 0.89–1.11 | 0.88–1.10 |
| | | Late collapse | SVI 2010, per 0.1 | 0.97 | 0.86–1.10 | 0.85–1.10 |
| | | Persistently constrained | SVI 2010, per 0.1 | 1.05 | 0.95–1.17 | 0.94–1.16 |
| | | Chronically low | SVI 2010, per 0.1 | 0.87 | 0.78–0.97 | 0.76–0.98 |
| | | Steady decline | Urban vs rural | 1.66 | 0.79–3.47 | 0.81–5.16 |
| | | Early collapse | Urban vs rural | 2.08 | 0.98–4.42 | 0.97–6.02 |
| | | Late collapse | Urban vs rural | 1.20 | 0.53–2.76 | 0.37–4.83 |
| | | Persistently constrained | Urban vs rural | 5.20 | 2.66–10.19 | 2.57–17.40 |
| | | Chronically low | Urban vs rural | 8.54 | 4.19–17.40 | 4.26–28.35 |
| Linear | SVI 2022 and urbanicity (n = 816) | Steady decline | SVI 2022, per 0.1 | 1.25 | 1.10–1.43 | 1.11–1.46 |
| | | Early collapse | SVI 2022, per 0.1 | 1.14 | 0.99–1.30 | 1.00–1.32 |
| | | Late collapse | SVI 2022, per 0.1 | 1.03 | 0.89–1.19 | 0.88–1.23 |
| | | Persistently constrained | SVI 2022, per 0.1 | 1.27 | 1.13–1.44 | 1.16–1.46 |
| | | Chronically low | SVI 2022, per 0.1 | 1.07 | 0.94–1.21 | 0.94–1.24 |
| | | Steady decline | Urban vs rural | 1.45 | 0.60–3.53 | 0.62–6.15 |
| | | Early collapse | Urban vs rural | 1.11 | 0.44–2.79 | 0.53–3.52 |
| | | Late collapse | Urban vs rural | 1.43 | 0.53–3.87 | 0.49–7.24 |
| | | Persistently constrained | Urban vs rural | 3.15 | 1.40–7.09 | 1.38–13.01 |
| | | Chronically low | Urban vs rural | 9.19 | 4.06–20.77 | 4.28–40.05 |
| Linear | SVI components and urbanicity (n = 807) | Steady decline | Black, per 10 percentage points | 1.03 | 0.87–1.22 | 0.87–1.22 |
| | | Early collapse | Black, per 10 percentage points | 1.04 | 0.88–1.24 | 0.88–1.26 |
| | | Late collapse | Black, per 10 percentage points | 1.02 | 0.85–1.24 | 0.82–1.26 |
| | | Persistently constrained | Black, per 10 percentage points | 1.00 | 0.85–1.16 | 0.83–1.17 |
| | | Chronically low | Black, per 10 percentage points | 1.01 | 0.86–1.18 | 0.81–1.20 |
| | | Steady decline | Poverty, per 10 percentage points | 1.18 | 0.72–1.91 | 0.75–1.88 |
| | | Early collapse | Poverty, per 10 percentage points | 1.23 | 0.75–2.03 | 0.75–2.03 |
| | | Late collapse | Poverty, per 10 percentage points | 1.00 | 0.57–1.76 | 0.55–1.81 |
| | | Persistently constrained | Poverty, per 10 percentage points | 1.22 | 0.78–1.92 | 0.82–1.97 |
| | | Chronically low | Poverty, per 10 percentage points | 1.21 | 0.76–1.92 | 0.82–1.89 |
| | | Steady decline | Median income, per US$10,000 | 0.77 | 0.59–1.00 | 0.60–0.98 |
| | | Early collapse | Median income, per US$10,000 | 1.00 | 0.79–1.26 | 0.78–1.33 |
| | | Late collapse | Median income, per US$10,000 | 0.97 | 0.75–1.25 | 0.70–1.21 |

| Model | Specification | Class | Term | RRR | Model-based 95% CI | Cluster-bootstrap 95% CI |
|---|---|---|---|---|---|---|
| | | Persistently constrained | Median income, per US$10,000 | 0.89 | 0.72–1.10 | 0.74–1.10 |
| | | Chronically low | Median income, per US$10,000 | 1.06 | 0.86–1.31 | 0.82–1.40 |
| | | Steady decline | Urban vs rural | 1.36 | 0.54–3.42 | 0.56–7.81 |
| | | Early collapse | Urban vs rural | 1.06 | 0.41–2.76 | 0.57–4.33 |
| | | Late collapse | Urban vs rural | 1.41 | 0.51–3.92 | 0.52–9.99 |
| | | Persistently constrained | Urban vs rural | 3.60 | 1.57–8.27 | 1.58–20.45 |
| | | Chronically low | Urban vs rural | 9.00 | 3.88–20.86 | 4.17–52.09 |
| Linear | SVI 2010 and urbanicity (n = 817) | Steady decline | SVI 2010, per 0.1 | 1.22 | 1.07–1.39 | 1.05–1.42 |
| | | Early collapse | SVI 2010, per 0.1 | 1.09 | 0.96–1.25 | 0.96–1.25 |
| | | Late collapse | SVI 2010, per 0.1 | 0.97 | 0.84–1.13 | 0.84–1.13 |
| | | Persistently constrained | SVI 2010, per 0.1 | 1.15 | 1.02–1.30 | 1.05–1.28 |
| | | Chronically low | SVI 2010, per 0.1 | 0.98 | 0.87–1.11 | 0.87–1.12 |
| | | Steady decline | Urban vs rural | 1.65 | 0.68–3.96 | 0.68–7.13 |
| | | Early collapse | Urban vs rural | 1.20 | 0.48–3.00 | 0.57–4.01 |
| | | Late collapse | Urban vs rural | 1.49 | 0.55–3.99 | 0.45–7.77 |
| | | Persistently constrained | Urban vs rural | 3.78 | 1.69–8.42 | 1.71–16.31 |
| | | Chronically low | Urban vs rural | 9.88 | 4.39–22.23 | 4.67–44.37 |

*Note:* Relative risk ratios (RRR) relative to the stable-adequate class. Cluster-bootstrap intervals are percentile intervals from 1,000 resamples of Mississippi's 82 counties. Urban, population-weighted tract centroid in a 2020 Census urban area. SVI, CDC/ATSDR Social Vulnerability Index (overall percentile, state-ranked); SVI 2010 mapped to 2020 tracts by shared land area. Associations are cross-sectional. Source: Data Axle; CDC/ATSDR SVI 2010 and 2022; American Community Survey 2018–2022; US Census Bureau.

**Table S11.** Spatial clustering of trajectory classes and index values.

| Class | Spline link: join-count z | P | Linear: join-count z | P |
|---|---|---|---|---|
| Stable adequate | 2.62 | 0.004 | 1.55 | 0.060 |
| Steady decline | 3.18 | <0.001 | 3.35 | <0.001 |
| Early collapse | 0.55 | 0.290 | 1.07 | 0.143 |
| Late collapse | 1.64 | 0.050 | 0.97 | 0.166 |
| Persistently constrained | −0.96 | 0.831 | −1.44 | 0.925 |
| Chronically low | 4.55 | <0.001 | 5.68 | <0.001 |

*Note:* Join-count standard deviates test whether tracts of the same class adjoin more often than expected under spatial randomness (first-order queen contiguity, 2020 tracts); positive values indicate clustering. Getis-Ord Gi* on the 2024 dollar-aware index (queen contiguity including each tract) identified 49 significant high-adequacy tracts (24 at the 95% level and 25 at the 99% level), no significant low-adequacy clusters and 757 tracts that were not significant: still-served towns form local clusters, whereas low adequacy is the statewide background rather than a local deviation below the mean. Source: Data Axle; US Census Bureau TIGER/Line 2022.

#### Supplementary Figures

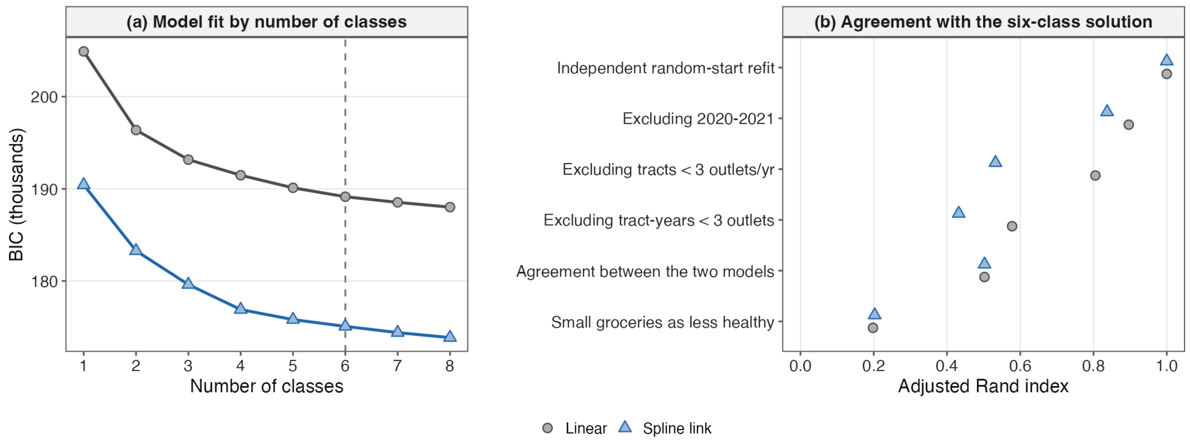


**Figure S1.** Data coverage and where the two indices differ. (a) All Mississippi establishment records in the annual Data Axle files; shaded, database-wide coverage expansions (2001; 2012–2013). (b, c) Mean difference between the standard and dollar-aware mRFEI in tract-years where dollar stores and healthy outlets coexist, (b) by 2022 Social Vulnerability Index quartile and (c) by number of dollar stores in the tract, pooled 1997–2024 and in 2024. Source: Data Axle; CDC/ATSDR SVI 2022.

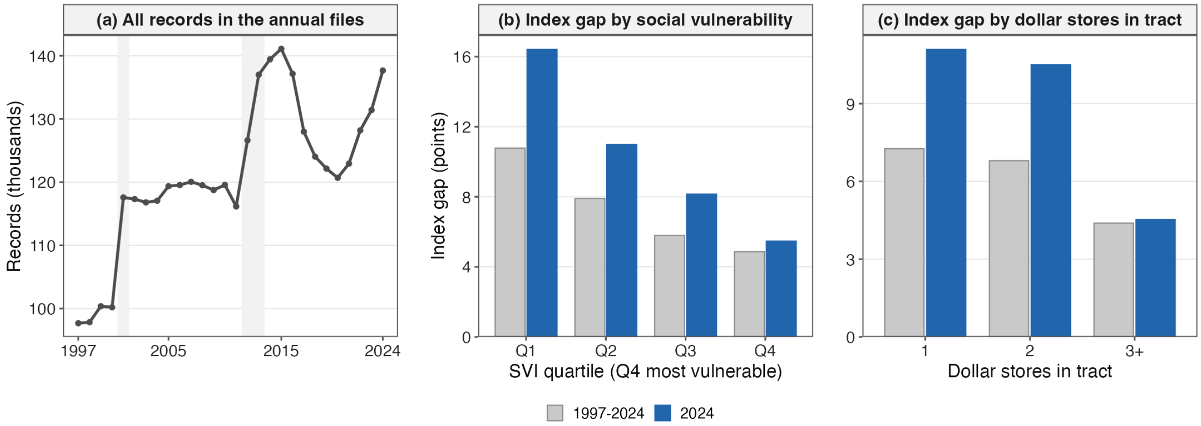


**Figure S2.** Model selection and stability of the six-class solution, spline-link and linear latent class models (817 tracts). (a) Bayesian Information Criterion (BIC) for one to eight classes; dashed line, the selected six-class solution. (b) Agreement (adjusted Rand index) between the main six-class solution and the solution from each sensitivity analysis: an independent refit from new random starts; excluding 2020–2021; excluding tracts averaging fewer than three food outlets per year; excluding tract-years with fewer than three outlets; counting grocery stores with three or fewer employees as less healthy; and agreement between the two specifications. Source: Data Axle.

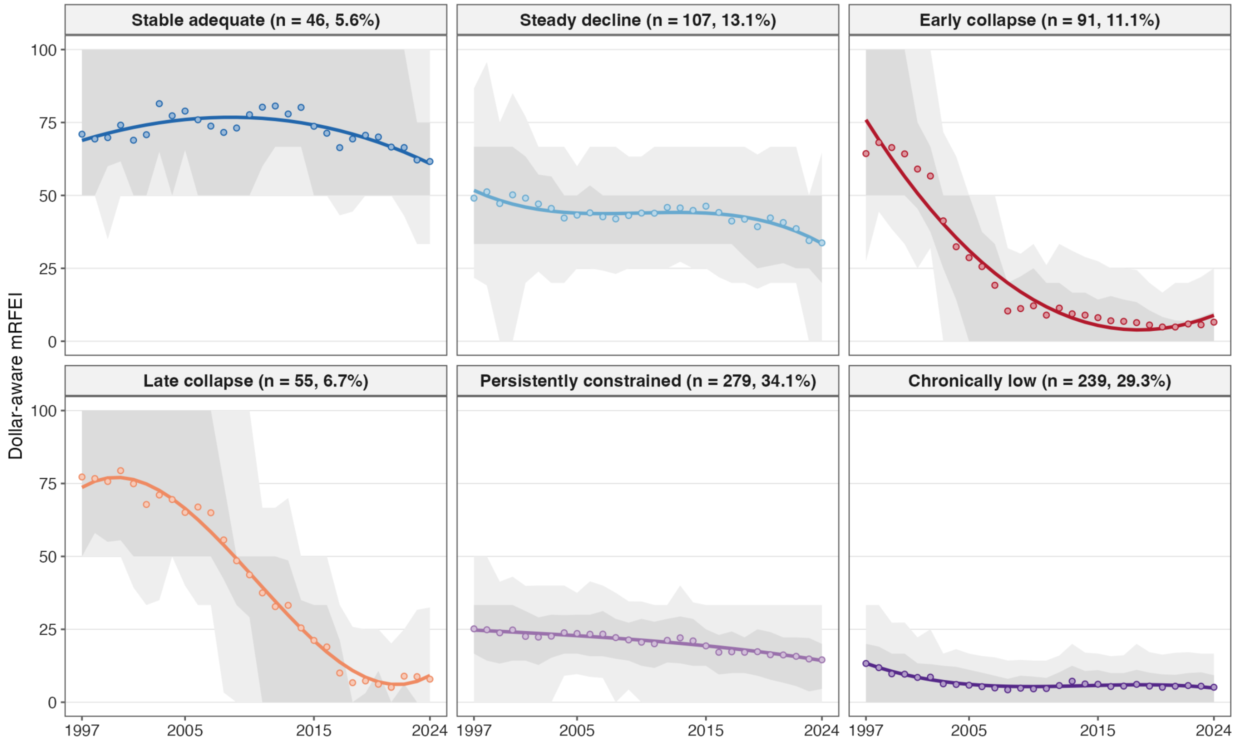


**Figure S3.** Trajectories of the dollar-aware mRFEI, 1997–2024, for six classes of Mississippi census tracts under the linear latent class model (sensitivity specification; 817 tracts). Grey bands, 10th–90th (light) and 25th–75th (dark) percentiles of tract values in each year; points, observed class mean; coloured line and band, model-predicted class trajectory with 95% interval. Years with no food retailer are treated as missing. Source: Data Axle.

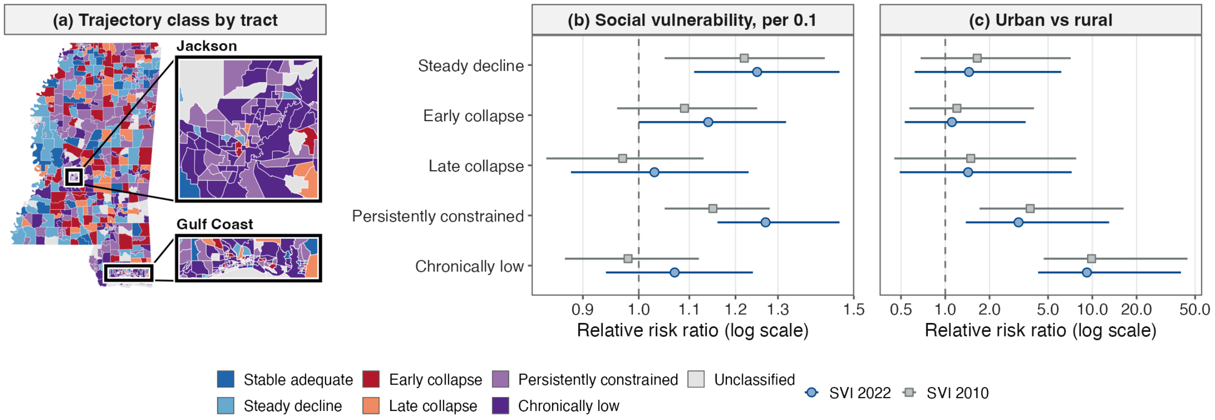


**Figure S4.** Geography and tract correlates of the trajectory classes under the linear latent class model (sensitivity specification). (a) Class of each Mississippi census tract, 1997–2024 (2020 boundaries), with the Jackson metropolitan area and the Gulf Coast enlarged at right; unclassified, 61 tracts with a food retailer in fewer than 15 of 28 years. (b, c) Relative risk ratios from multinomial models with stable adequate as reference, for (b) social vulnerability (per 0.1) and (c) urban location, using 2022 SVI or 2010 SVI mapped to 2020 tracts; bars, 95% intervals from a county cluster bootstrap (1,000 replicates). Associations are cross-sectional. Source: Data Axle; CDC/ATSDR SVI 2010 and 2022; US Census Bureau TIGER/Line 2022.
