## additional file 2 for "A dollar-aware food-environment index and a 27-year trajectory typology: a measurement foundation for diet and childhood-obesity research in Mississippi, 1997–2024"

 doi:10.64898/2026.08.20.26360912

### Additional file 2

#### STROBE-nut checklist

Reference: Lachat C, Hawwash D, Ocké MC, et al. Strengthening the Reporting of Observational Studies in Epidemiology – Nutritional Epidemiology (STROBE-nut): an extension of the STROBE statement. PLoS Med. 2016;13(6):e1002036.

This is an observational study of the retail food environment: annual establishment records for all 878 Mississippi census tracts, 1997–2024, summarised as a food-environment index, latent trajectory classes and their cross-sectional associations with tract characteristics. The analytic units are establishments and census tracts; the study does not include individual dietary assessment or nutritional-status measurement. The core STROBE items therefore apply throughout, whereas most STROBE-nut extensions on dietary assessment are not applicable (n/a), as noted for each item. Locations refer to section, table and figure numbers in the revised manuscript; the trajectory analysis is additionally reported against the GRoLTS checklist (Additional file 3).

| Section / topic | Item | STROBE recommendation | STROBE-nut extension | Reported in |
|---|---|---|---|---|
| Title and abstract | 1 | (a) Indicate the study's design with a commonly used term in the title or the abstract. (b) Provide in the abstract an informative and balanced summary of what was done and what was found. | nut-1: State the dietary/nutritional assessment method(s) used in the title, abstract or keywords. | Title; Abstract (Methods, Results). nut-1: n/a (no dietary assessment) |
| **Introduction** | | | | |
| Background/ rationale | 2 | Explain the scientific background and rationale for the investigation being reported. | — | Section 1 (Background) |
| Objectives | 3 | State specific objectives, including any prespecified hypotheses. | — | Section 1, final paragraph (four aims) |
| **Methods** | | | | |
| Study design | 4 | Present key elements of study design early in the paper. | — | Abstract (Methods); Sections 2.1 and 2.5 |
| Setting | 5 | Describe the setting, locations and relevant dates, including periods of recruitment, exposure, follow-up and data collection. | nut-5: Describe any characteristics of the study settings that might affect the dietary intake or nutritional status of the participants, if applicable. | Section 1 (Mississippi context); Section 2.1 (annual files, 1997–2024; 878 tracts). nut-5: n/a |

| Section / topic | Item | STROBE recommendation | STROBE-nut extension | Reported in |
|---|---|---|---|---|
| Participants (units of analysis) | 6 | Give the eligibility criteria and the sources and methods of selection of participants. | nut-6: Report particular dietary, physiological or nutritional characteristics that were considered when selecting the target population. | Sections 2.1 and 2.5 (all 878 tracts; 817 modelled tracts with a food retailer in $\geq$15 of 28 years). nut-6: n/a |
| Variables | 7 | Clearly define all outcomes, exposures, predictors, potential confounders and effect modifiers. Give diagnostic criteria, if applicable. | nut-7.1: Clearly define foods, food groups, nutrients or other food components. nut-7.2: When using dietary patterns or indices, describe the methods to obtain them and their nutritional properties. | Sections 2.2 (retail categories), 2.3 (indices), 2.4 (SVI, ACS, urbanicity), 2.5 (trajectory classes). nut-7.1: outlet categories defined (Section 2.2); nut-7.2: n/a (food-environment index, not a dietary index; Section 2.3) |
| Data sources/ measurement | 8 | For each variable of interest, give sources of data and details of methods of assessment (measurement). Describe comparability of assessment methods if there is more than one group. | nut-8.1–8.6: Dietary assessment method; food composition data; nutrient requirements; biomarkers; assessment of non-dietary data; validity of dietary or nutritional assessment. | Sections 2.1–2.4; Additional file 1: Supplementary Methods S1–S2 and Tables S1–S3. nut-8.5: classification validated against company-reported counts and reviewed for misclassification (Section 2.2); other nut-8 items n/a |
| Bias | 9 | Describe any efforts to address potential sources of bias. | nut-9: Report how bias in dietary or nutritional assessment was addressed, e.g. misreporting, changes in habits as a result of being measured, or data imputation from other sources. | Section 2.2 (validation; misclassification review); Section 2.5 (seven stability analyses; vendor coverage checks); Section 3.1. nut-9: n/a |
| Study size | 10 | Explain how the study size was arrived at. | — | Sections 2.1 and 2.5 (complete establishment census; all Mississippi tracts) |
| Quantitative variables | 11 | Explain how quantitative variables were handled in the analyses. If applicable, describe which groupings were chosen and why. | nut-11: Explain categorisation of dietary/nutritional data and the choice of reference categories, if applicable. | Sections 2.3–2.5 (index 0–100 with spline link; SVI per 0.1; components per 10 points and US$10,000; urban definition). nut-11: n/a |
| Statistical methods | 12 | (a) Describe all statistical methods, including those used to control for confounding. (b) Describe any methods used to examine subgroups and interactions. (c) Explain how missing data were addressed. (d) If applicable, describe analytical methods taking account of sampling strategy. (e) Describe any sensitivity analyses. | nut-12.1–12.3: Methods to combine dietary data; energy adjustment, intake modelling and weighting; adjustments for measurement error. | Section 2.5; Additional file 1: Supplementary Methods S3–S5. (c) Years with no food retailer treated as missing; 61 tracts described separately (Section 2.5). (d) County cluster bootstrap for spatial clustering. (e) Seven stability analyses (Section 2.5; Table 2). nut-12: n/a |
| **Results** | | | | |
| Participants (units of analysis) | 13 | (a) Report numbers of individuals at each stage of the study. (b) Give reasons for non-participation at each stage. (c) Consider use of a flow diagram. | nut-13: Report the number of individuals excluded based on missing, incomplete or implausible dietary/nutritional data. | Sections 2.5 and 3.3 (878 tracts; 817 modelled; 61 not modelled, 17 with no food retailer). nut-13: n/a |
| Descriptive data | 14 | (a) Give characteristics of study participants and information on exposures and potential confounders. (b) Indicate the number of participants with missing data for each variable of interest. | nut-14: Give the distribution of participant characteristics across the exposure variables if applicable; specify whether food consumption of the total population or consumers only were used. | Sections 3.1 and 3.3; Table 1; Additional file 1: Tables S4–S5 and S8. (b) Section 2.4 (SVI 2010 for 872 of 878 tracts); Table 3 note (n = 816 with valid SVI). nut-14: n/a |

| Section / topic | Item | STROBE recommendation | STROBE-nut extension | Reported in |
|---|---|---|---|---|
| Outcome data | 15 | Report numbers of outcome events or summary measures. | — | Sections 3.1–3.3; Figures 1–2; Table 1; Additional file 1: Tables S4 and S6 |
| Main results | 16 | (a) Give unadjusted estimates and, if applicable, confounder-adjusted estimates and their precision (e.g. 95% CI). Make clear which confounders were adjusted for and why. (b) Report category boundaries when continuous variables were categorised. (c) If relevant, consider translating estimates of relative risk into absolute risk. | nut-16: Specify if nutrient intakes are reported with or without inclusion of dietary supplement intake, if applicable. | Section 3.5; Table 3 (relative risk ratios with cluster-bootstrap and model-based 95% CI); Figure 3. (b) SVI quartiles and urban definition (Sections 2.4 and 3.2). nut-16: n/a |
| Other analyses | 17 | Report other analyses done, e.g. analyses of subgroups and interactions, and sensitivity analyses. | nut-17: Report any sensitivity analysis (e.g. exclusion of misreporters or outliers) and data imputation, if applicable. | Section 3.4 (Table 2); Section 3.5 (components model; SVI 2010); Section 3.6 (spatial structure); Additional file 1: Tables S3 and S7–S11. nut-17: non-dietary sensitivity analyses reported; dietary misreporting n/a |
| **Discussion** | | | | |
| Key results | 18 | Summarise key results with reference to study objectives. | — | Section 4, first paragraph |
| Limitations | 19 | Discuss limitations of the study, taking into account sources of potential bias or imprecision. Discuss both direction and magnitude of any potential bias. | nut-19: Describe the main limitations of the data sources and assessment methods used and implications for the interpretation of the findings. | Section 4.2. nut-19: limitations of the commercial business data, the outlet classification and its likely direction of bias, and the absence of dietary assessment (Section 4.2) |
| Interpretation | 20 | Give a cautious overall interpretation of results considering objectives, limitations, multiplicity of analyses, results from similar studies and other relevant evidence. | nut-20: Report the nutritional relevance of the findings, given the complexity of diet or nutrition as an exposure. | Sections 4 and 4.1; Section 5 (Conclusions). nut-20: relevance of the retail food environment to diet and diet-related disease (Sections 1 and 4) |
| Generalisability | 21 | Discuss the generalisability (external validity) of the study results. | — | Section 4, sixth paragraph |
| **Other information** | | | | |
| Funding | 22 | Give the source of funding and the role of the funders for the present study and, if applicable, for the original study on which the present article is based. | — | Declarations (Funding) |
| Ethics | — | — | nut-22.1: Describe the procedure for consent and study approval from ethics committee(s). | Declarations (Ethics approval and consent to participate: no human participants; IRB approval not required) |
| Supplementary material | — | — | nut-22.2: Provide data collection tools and data as online material or explain how they can be accessed. | Declarations (Availability of data and materials: licensed Data Axle records; analysis code available from the corresponding author on request); Additional file 1 |

*Note:* n/a, not applicable because no dietary intake or nutritional status was measured. GRoLTS, Guidelines for Reporting on Latent Trajectory Studies.
