## additional file 3 for "A dollar-aware food-environment index and a 27-year trajectory typology: a measurement foundation for diet and childhood-obesity research in Mississippi, 1997–2024"

 doi:10.64898/2026.08.20.26360912

### Additional file 3

#### GRoLTS checklist: Guidelines for Reporting on Latent Trajectory Studies

Reference: van de Schoot R, Sijbrandij M, Winter SD, Depaoli S, Vermunt JK. The GRoLTS-Checklist: guidelines for reporting on latent trajectory studies. Struct Equ Modeling. 2017;24(3):451–67.

The trajectory analysis models the annual dollar-aware modified Retail Food Environment Index (0–100) for 817 Mississippi census tracts, 1997–2024, with latent class mixed models in the lcmm package for R. The primary specification uses a monotone spline link; a linear (Gaussian) specification is reported as a sensitivity analysis. Locations refer to the revised manuscript and to Additional file 1.

| Item | Checklist question | Reported? | Where and how |
|---|---|---|---|
| 1 | Is the metric of time used in the statistical model reported? | Yes | Calendar year, 1997–2024, scaled as (year − 1997)/10 (Section 2.5; Additional file 1: Supplementary Methods S3) |
| 2 | Is information presented about the mean and variance of time within a wave? | Not applicable | Each wave is a calendar year of establishment records; every tract is observed on the same annual schedule, so time does not vary within a wave (Section 2.1) |
| 3a | Is the missing data mechanism reported? | Yes | The index is undefined in tract-years with no food retailer; these tract-years (3.7% in modelled tracts) are treated as missing, not as zero (Section 2.5) |
| 3b | Is a description provided of what variables are related to attrition or missing data? | Yes | Missingness arises in tracts with few or no food retailers; tracts with a retailer in fewer than 15 of 28 years (61 tracts) are described separately, including their retailer counts and number of children (Sections 2.5, 3.3 and 3.4) |
| 3c | Is a description provided of how missing data in the analyses were dealt with? | Yes | Maximum likelihood estimation using all available tract-years; sensitivity analysis treating tract-years with fewer than three retailers as missing (Section 2.5; Table 2; Additional file 1: Table S9) |
| 4 | Is information about the distribution of the observed variables included? | Yes | 30.4% of modelled tract-years at 0 and 5.0% at 100; median 5 food retailers per tract-year (interquartile range 3–10); percentile bands of tract values in each year (Sections 2.5 and 3.3; Figure 2; Additional file 1: Table S6) |
| 5 | Is the software mentioned? | Yes | R, lcmm package (functions lcmm with a spline link and hlme for the linear specification) (Section 2.5; Additional file 1: Supplementary Methods S3) |
| 6a | Are alternative specifications of within-class heterogeneity considered (e.g. latent class growth analysis versus growth mixture modelling) and clearly documented? | Yes | Latent class growth specification without within-class random effects, so that classes describe shared mean shapes; stated and justified in Section 2.5 |

| Item | Checklist question | Reported? | Where and how |
|---|---|---|---|
| 6b | Are alternative specifications of the between-class differences in the variance–covariance matrix structure considered and clearly documented? | Not applicable | No within-class random effects are estimated, so there is no class-specific variance–covariance matrix; the residual variance is common to all classes (Section 2.5; Additional file 1: Supplementary Methods S3) |
| 7 | Are alternative shape or functional forms of the trajectories described? | Yes | Cubic class-specific trajectories; link functions compared: monotone spline (five equidistant knots, primary), linear (sensitivity) and beta (did not converge) (Section 2.5; Additional file 1: Table S7) |
| 8 | If covariates have been used, can analyses still be replicated? | Yes | No covariates enter the trajectory models; tract characteristics are related to class membership afterwards in multinomial models, fully specified with units and inference (Section 2.5; Table 3; Additional file 1: Table S10) |
| 9 | Is information reported about the number of random start values and final iterations included? | Yes | 24 random starts for each number of classes; the best start iterated to convergence; an independent refit from new random starts reproduced the six-class solution (ARI 1.00) (Sections 2.5 and 3.4; Additional file 1: Supplementary Methods S3) |
| 10 | Are the model comparison (and selection) tools described from a statistical perspective? | Yes | BIC, sample-size-adjusted BIC, relative entropy, average posterior probability of assignment, occupancy odds ratio, smallest class size, stability across adjacent solutions and substantive distinctness (Section 2.5; Section 3.3; Additional file 1: Table S7) |
| 11 | Are the total number of fitted models reported, including a one-class solution? | Yes | One to eight classes for both the spline-link and linear specifications (16 models) (Additional file 1: Table S7; Figure S2a) |
| 12 | Are the number of cases per class reported for each model (absolute sample size, or proportion)? | Yes | Smallest class size for every model (Additional file 1: Table S7); full class sizes for the six-class solutions (Table 1; Additional file 1: Table S8); class composition at five, seven and eight classes described (Section 3.3) |
| 13 | If classification of cases in a trajectory is the goal, is entropy reported? | Yes | Relative entropy for every model (Additional file 1: Table S7); 0.964 for the selected spline-link solution, with class-specific APPA 0.953–0.983 (Section 3.3; Additional file 1: Table S8) |
| 14a | Is a plot included with the estimated mean trajectories of the final solution? | Yes | Figure 2 (spline link) and Additional file 1: Figure S3 (linear) |
| 14b | Are plots included with the estimated mean trajectories for each model? | Partly | Plots are shown for the six-class solutions of both specifications; the class structure of the five-, seven- and eight-class solutions is described in the text (Section 3.3; Additional file 1: Table S7) |
| 14c | Is a plot included of the combination of estimated means of the final model and the observed individual trajectories split out for each latent class? | Yes | Figure 2 shows, for each class, the model-predicted trajectory with its 95% interval, the observed class mean and the 10th–90th and 25th–75th percentile bands of observed tract values (bands are used in place of individual lines for legibility across 817 tracts) |
| 15 | Are characteristic descriptive measures of the final class solution reported (means, SD, n, CI, etc.)? | Yes | Tracts and children per class, observed and predicted index values, social and urban characteristics and distance to grocery stores (Table 1; Additional file 1: Table S8); model fit by class (Section 3.3) |
| 16 | Are the syntax files available (either in the appendix, supplementary materials, or from the authors)? | Yes | Analysis code is available from the corresponding author on request (Declarations: Availability of data and materials); model specification in Additional file 1: Supplementary Methods S3 |

*Note:* APPA, average posterior probability of assignment; ARI, adjusted Rand index; BIC, Bayesian Information Criterion.
